# Risk factors for breast cancer subtypes by race and ethnicity: A scoping review of the literature

**DOI:** 10.1101/2024.03.18.24304210

**Authors:** Amber N Hurson, Thomas U Ahearn, Hela Koka, Brittany D Jenkins, Alexandra R Harris, Sylvia Roberts, Sharon Fan, Jamirra Franklin, Gisela Butera, Renske Keeman, Audrey Y Jung, Pooja Middha, Gretchen L Gierach, Xiaohong R Yang, Jenny Chang-Claude, Rulla M Tamimi, Melissa A Troester, Elisa V Bandera, Mustapha Abubakar, Marjanka K Schmidt, Montserrat Garcia- Closas

## Abstract

**Background:** Breast cancer is comprised of distinct molecular subtypes. Studies have reported differences in risk factor associations with breast cancer subtypes, especially by tumor estrogen receptor (ER) status, but their consistency across racial and ethnic populations has not been comprehensively evaluated.

**Methods:** We conducted a qualitative, scoping literature review using the Preferred Reporting Items for Systematic Reviews and Meta-analysis, extension for Scoping Reviews to investigate consistencies in associations between 18 breast cancer risk factors (reproductive, anthropometric, lifestyle, and medical history) and risk of ER-defined subtypes in women who self-identify as Asian, Black or African American, Hispanic or Latina, or White. We reviewed publications between January 1, 1990 and July 1, 2022. Etiologic heterogeneity evidence (convincing, suggestive, none, or inconclusive) was determined by expert consensus.

**Results:** Publications per risk factor ranged from 14 (benign breast disease history) to 66 (parity). Publications were most abundant for White women, followed by Asian, Black or African American, and Hispanic or Latina women. Etiologic heterogeneity evidence was strongest for parity, followed by age at first birth, post-menopausal BMI, oral contraceptive use, and estrogen-only and combined menopausal hormone therapy. Evidence was limited for other risk factors. Findings were consistent across racial and ethnic groups, although the strength of evidence varied.

**Conclusion:** The literature supports etiologic heterogeneity by ER for some established risk factors that are consistent across race and ethnicity groups. However, in non-White populations evidence is limited. Larger, more comparable data in diverse populations is needed to better characterize breast cancer etiologic heterogeneity.

## INTRODUCTION

Globally, female breast cancer was the most frequently diagnosed cancer (11.7% of all cancer diagnoses, over 2.2 million new cases) and the leading cause of cancer-related deaths (15.5% of all cancer deaths) among women in 2020.^1^ In the US, about 80% of breast cancers are estrogen receptor (ER)-positive, of which about 70% are the luminal A subtype (defined herein as breast cancers that are ER and/or progesterone receptor (PR)-positive, and human epidermal growth factor receptor 2 (HER2)-negative). The remaining are ER-negative, of which about 10% are triple negative (defined as tumors that are negative for ER, PR, and HER2).^2^ Incidence of breast cancer subtypes in the US varies by race and ethnicity, with incidence of luminal A highest in Non-Hispanic White women and triple negative highest in Non-Hispanic Black women.^2–4^

An expanding body of literature, including many reviews and pooled analyses, indicates distinct etiologies exist for some breast cancer subtypes, with some risk factors having unique associations with risk of particular subtypes. ^5–11^ For example, some studies have found higher levels of parity to be associated with a lower risk of luminal A disease, but a higher risk of triple negative disease.^5,7^ Similarly, postmenopausal obesity and the use of menopausal hormone therapy (MHT) have been shown to be more strongly associated with elevated risks of luminal than non-luminal breast cancers.^12–15^

Given the differences in the incidence patterns, biological behavior, and clinical presentation of breast cancer subtypes, an improved understanding of etiologic heterogeneity is crucial for characterizing disease burden and informing public health and research priorities. Despite growing interest in this subject, significant gaps remain. Most studies and reviews have focused on self-reported White populations. While some individual and pooled studies have suggested differences in subtype associations with certain risk factors across racially and ethnically diverse populations,^6,16–18^ most reviews have not directly compared these associations across racial and ethnic populations. Clarifying whether the associations between risk factors and risk of disease subtypes are consistent across racial and ethnic populations has important implications for understanding disease etiology and informing targeted prevention strategies.

We performed a systematic scoping review of the literature to assess the current evidence on the associations between multiple breast cancer risk factors and risk of breast cancer subtypes defined primarily by tumor ER or triple negative status in Asian, Black or African American, Hispanic or Latina, and White women. The focus of this review is to evaluate heterogeneity in associations across these subtypes, as well as to evaluate the consistency of heterogeneity patterns across populations. In performing this review, we sought to provide a comprehensive evaluation of current data, and to highlight gaps in the literature in need of additional research.

## METHODS

We conducted a scoping review to systematically chart available literature on this topic, synthesize findings, and identify key gaps. The scoping review followed the methodological framework developed by Arksey and O’Malley, refined by Levac et al., and further revised by the Joanna Briggs Institute Manual for Evidence Synthesis.^19–21^ The framework stages of the review include (1) identifying the research question; (2) identifying the relevant studies; (3) study selection; (4) charting the data; and (5) collating, summarizing, and reporting the results. Our research protocol was registered with the Open Science Framework (OSF) (DOI: 10.17605/OSF.IO/TNG7K). The review was performed according to the Preferred Reporting Items for Systematic Reviews and Meta-analysis, extension for Scoping Reviews (PRISMA-ScR).^22^

### Literature Search

Literature search strategies were developed using medical subject headings (MeSH) and text words related to breast cancer, established breast cancer risk factors, and disease subtypes. The established breast cancer risk factors were grouped into the following categories: reproductive factors, anthropometric factors, lifestyle factors, and medical factors/medication use. The search strategy was developed by a review member (TUA) and further refined by a biomedical librarian (GB) using an iterative process. The following databases were searched for articles published from January 1, 1990 through July 1, 2022: PubMed (National Library of Medicine), Embase (Elsevier), CINAHL Plus (Cumulative Index to Nursing and Allied Health Literature - EBSCOhost), Web of Science: Core Collection (Clarivate Analytics), and Scopus (Elsevier). The final search strategy can be found in **Table S1**.

Eligible articles were required to have provided estimates of associations (e.g., odds ratios), between risk factors of interest and risk of breast cancer subtypes (defined by ER status, hormone receptor status, luminal status, or TN-status) by racial and ethnic groups. Results must have been generated from individual-level data, including studies that pooled individual-level data from multiple study populations. We broadly categorized race and ethnicity as Asian, Black or African American, Hispanic or Latina, and White women. We extracted results into these groups based on how the reviewed studies collected and reported race and ethnicity in their study. Included studies must have reported their results stratified by race and/or ethnicity or have generated their results from a study population that was reported to be at least 80% comprised of participants that reported to belong to one of these racial or ethnic groups. Studies that did not report racial and ethnic distributions were reviewed by the authors to determine if the study population could be reasonably assumed to be at least 80% comprised of Asian, Black or African American, Hispanic or Latina, or White women based on the country in which the study was performed. In analyzing these studies, we considered race and ethnicity as social constructs, acknowledging that many behavioral, cultural, lifestyle, and community- or neighborhood-level factors may differ across populations. These social factors may either have limited effect or play out differently across populations. Animal and cell-line studies, non-English articles, and abstracts with no access to the full text were excluded from the review.

### Study Selection

After articles were identified through the database searches, duplicates were removed using EndNote 20 (Clarivate 2020) reference manager software and results were imported into Covidence (www.covidence.org), a web-based screening collaboration software platform that streamlines the production of evidence synthesis and other literature reviews. A two-stage review process was used for study selection: title/abstract screening and full-text screening. At both stages, each article was screened by two out of seven possible reviewers in accordance with inclusion and exclusion criteria. Studies ruled ineligible by both reviewers were excluded, while those with discordant decisions on eligibility were arbitrated by a third screener. Interrater agreement was 95% on average (ranged from 82% to 100%). The screening processes and data extraction forms were piloted and calibrated by the screeners using a random sample of 25 articles, during which necessary changes to the screening procedures were made. **Figure S1** details the identification and screening of relevant publications.

### Data Extraction and Charting

Relevant data was extracted into a standardized data collection form using Covidence. As the cut points for numerical variables and the category definitions for ordinal variables varied across articles, we charted the effect estimate for the highest category in reference to the lowest category. Estimates were reparametrized, when required, to achieve this. Stratified effect estimates (e.g., by age, menopausal status) were meta-analyzed and charted as a single effect estimate. Tumor subtypes were categorized as either ER-positive or ER-negative. Other subtypes were not included in this review as evidence for etiologic heterogeneity by diverse populations is limited beyond the subtypes that are considered here.

When associations with risk of the ER-positive subtype were not reported, we allowed for associations with risk of subtypes defined as hormone receptor positive, luminal, luminal A, or ER-positive & PR-positive. For the ER-negative subtype group, associations with risk of triple negative or basal-like breast cancer were preferred, when available, as this subtype is the most etiologically distinct from ER-positive. When associations with risk of triple negative/basal-like or ER-negative subtype were not reported, we allowed subtypes defined as hormone receptor negative or ER-negative & PR-negative. When multiple eligible publications were published using a given study population, we extracted the most recently published estimates of association from the given study population for each risk factor. Published estimates from case-control and case-case study designs were charted separately. Plots of the extracted estimates were generated using the R software (version 4.2.0 http://cran.r-project.org/).

Evidence of etiologic heterogeneity was discussed amongst all coauthors, with final determinations made by a panel of breast cancer experts (ANH, TUA, RK, XRY, JCC, RMT, MAT, EVB, MA, MKS, MGC). Within each racial and ethnic group, each expert independently proposed whether the published evidence supported the presence of subtype heterogeneity (i.e., difference in direction or magnitude of association) as either convincing, suggestive, inconclusive, or no evidence of heterogeneity in risk by subtype heterogeneity (**Figure S2**). Race and ethnicity population groups with fewer than two published estimates for a given risk factor were not evaluated for heterogeneity by subtype. The group met to discuss conclusions and reach consensus.

## RESULTS

### Study Characteristics

A total of 6,517 articles were imported into Covidence from the five databases, of which 518 passed the screening eligibility criteria (see Methods) (**Figure S1**). Of these, 299 (58%) were excluded for various reasons, including not stratifying estimates by tumor subtype (n=63), not stratifying estimates by race and ethnicity (n=60), not reporting an estimate of effect (n=52), and other reasons (**Figure S1**). Finally, a total of 219 articles, representing 135 different study populations, were included in the review.

The numbers of the eligible articles for each risk factor are shown in **Table 1**. Most publications reported estimates for self-reported White women, followed by Asian, Black or African American, and Hispanic or Latina women. The proportion of published estimates that represented populations of self-reported White women ranged from 53% for breastfeeding to 90% for history of benign breast disease. The number of publications per risk factor ranged from n=14 for history of benign breast disease to n=66 for parity.

**Table 1.**
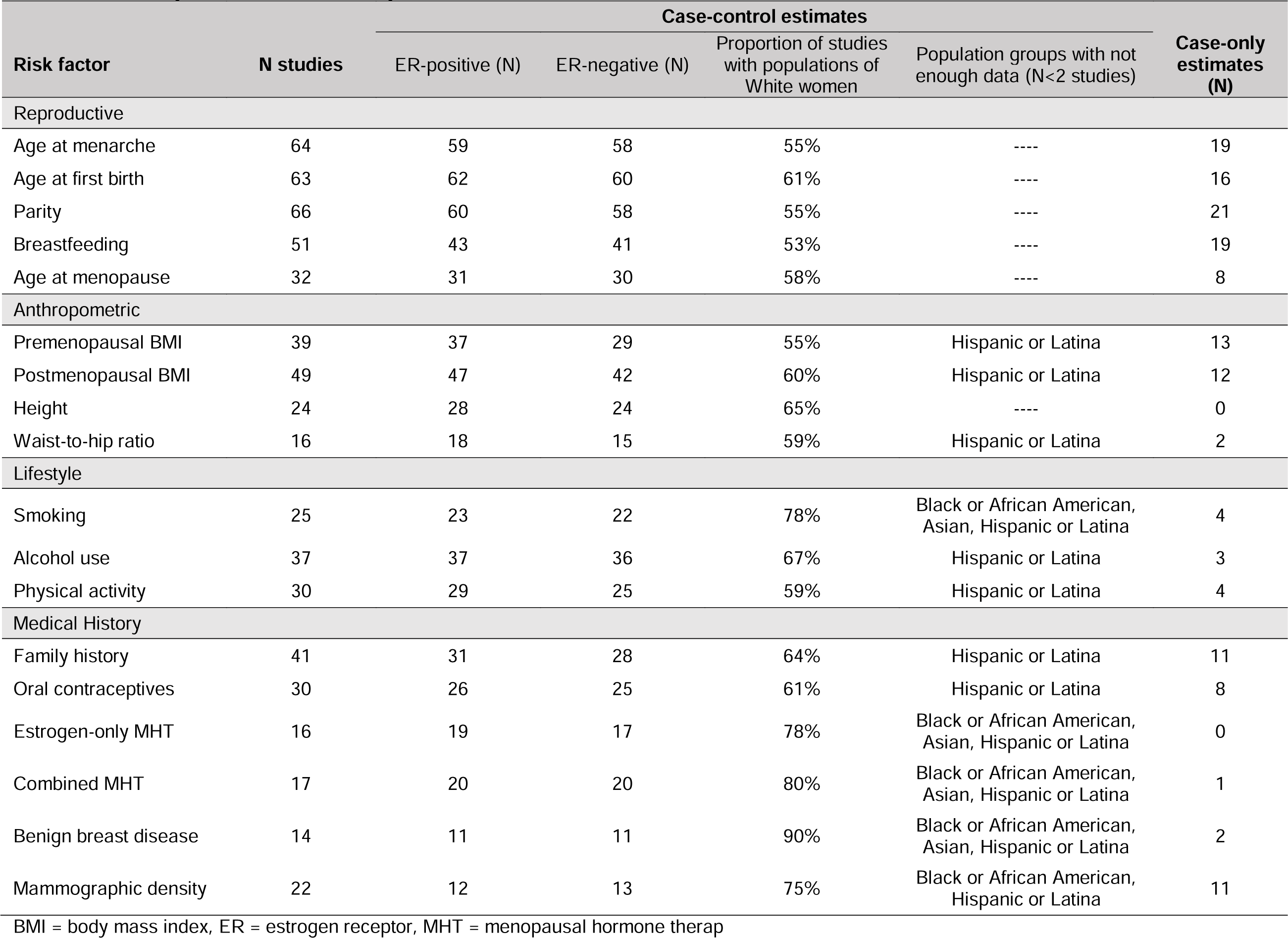
Summary of extracted articles by breast cancer risk factor.

An overview of all analyses and evidence for heterogeneity by ER status is shown in **Figure 1**.

**Figure 1.**
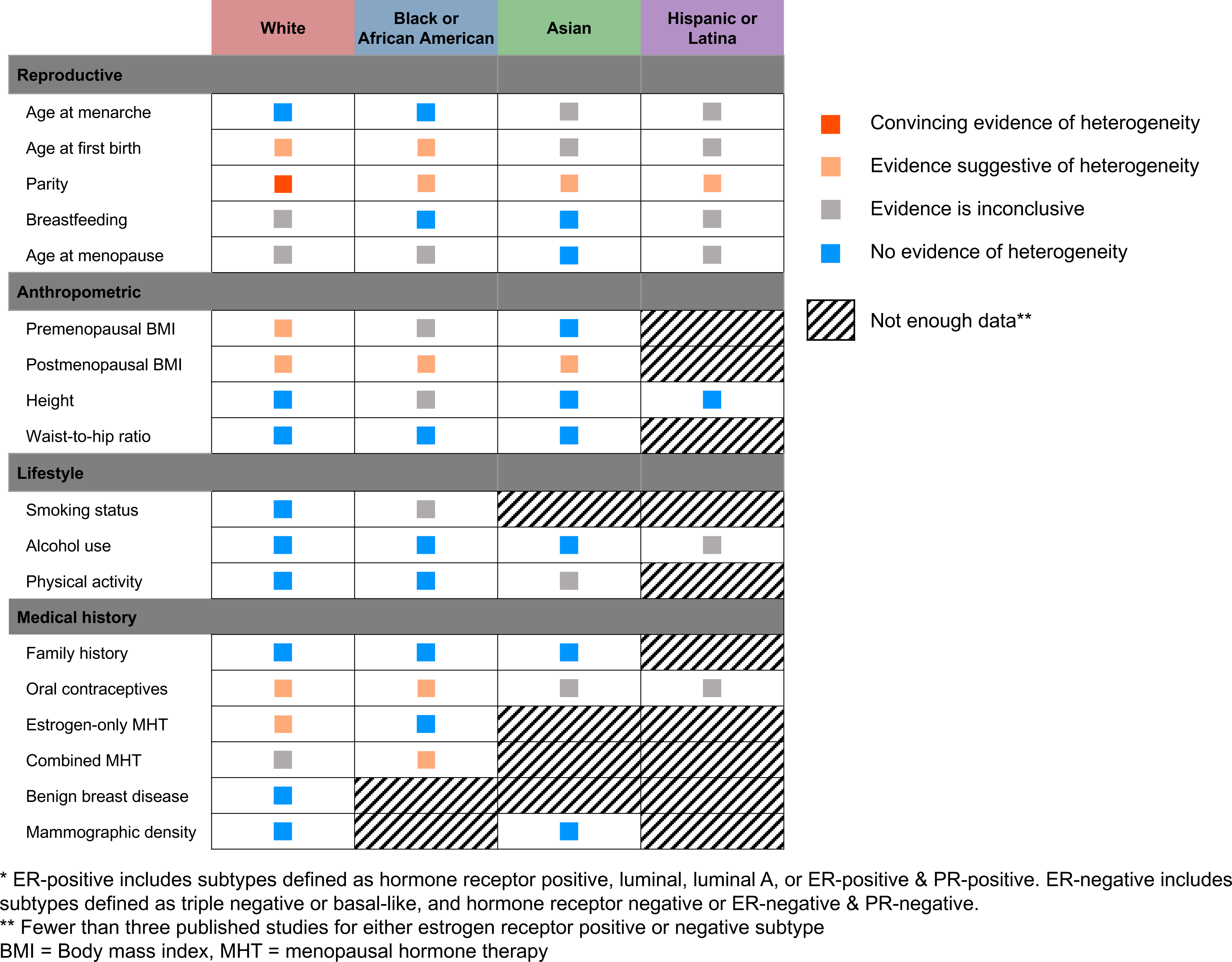
Summary of evidence for heterogeneity of associations with risk for breast cancer by estrogen receptor (ER) tumor status* among diverse racial and ethnic populations.

### Reproductive Factors

Of all 219 publications, there were 66 that reported effect estimates for parity, 64 for age at menarche, 63 for age at first birth, 51 for breastfeeding, and 32 for age at menopause. The published evidence of heterogeneity in subtype-specific risk associated with parity was judged to be convincing for White women and suggestive across the other race and ethnicity groups. For age at first birth, the evidence of heterogeneity was suggestive for Black or African American and White women. In all investigated populations, higher parity was more strongly associated with reduced risk of ER-positive than ER-negative disease (**Figure 2, Table S2**). Similarly, in populations of White and Black or African American women, later age at first birth was also more strongly associated with a higher risk of ER-positive than ER-negative disease; however, the evidence of heterogeneity in risk by subtype associated with age at first birth was inconclusive among populations of Asian and Hispanic or Latina populations because of high variability across studies and too few published studies, respectively (**Figure 3, Table S3**).

**Figure 2.**
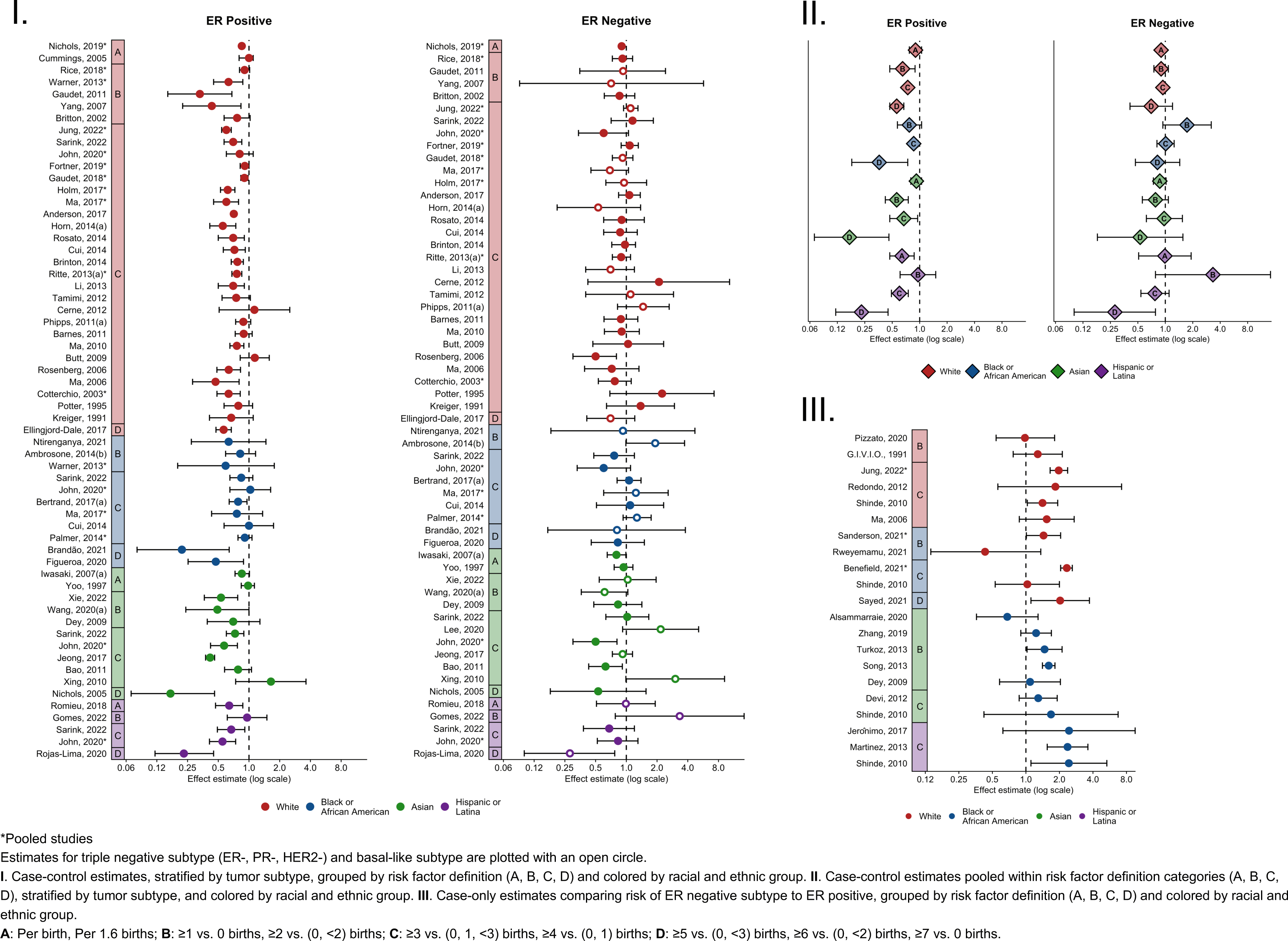
Published estimates for the effect of parity on breast cancer risk by tumor subtype and racial and ethnic group.

**Figure 3.**
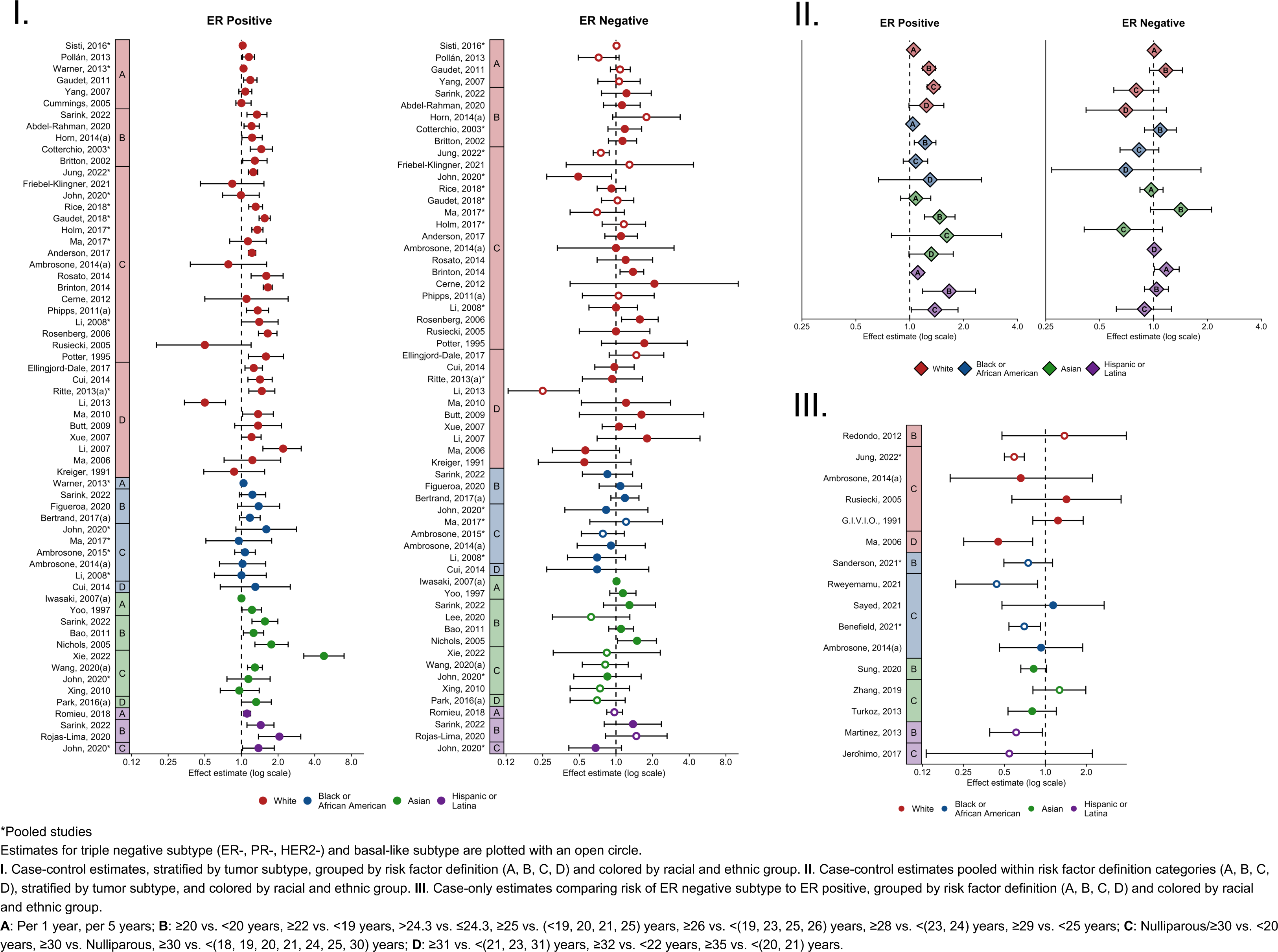
Published estimates for the effect of age at first birth on breast cancer risk by tumor subtype and racial and ethnic group.

For other reproductive factors—age at menarche, breastfeeding, and age at menopause—there was either inconclusive or no evidence of heterogeneity in risk by subtype for each race and ethnicity group (**Figures S3-S5, Tables S4-S6**). Later age at menarche was associated with a lower risk for both ER-positive and ER-negative disease, while breastfeeding and later age at menopause were respectively associated with a lower and higher risk of breast cancer. For breastfeeding, the published evidence among populations of White and Hispanic or Latina women was inconclusive as to whether the magnitude of the associations varies by subtype; and similarly, for age at menopause, among White, Black or African American, and Hispanic or Latina women, the evidence is inconclusive on whether the magnitude of the associations varies by subtype.

### Anthropometric Factors

Among the eligible publications, 49 reported effect estimates for postmenopausal BMI, 39 for premenopausal BMI, 24 for height, and 16 for waist-to-hip ratio. The evidence suggests heterogeneity in risk by subtype for pre- and post-menopausal BMI (**Figures 4-5, Tables S7-S8**), while the published associations with height and waist-to-hip ratio do not indicate heterogeneity by ER status (**Figures S6-S7, Tables S9-S10**). The evidence suggests higher postmenopausal BMI is more strongly associated with a higher risk of ER-positive disease than with ER-negative. This pattern was consistent across population groups, although there were too few studies in populations of Hispanic or Latina descent to be conclusive. For pre-menopausal BMI, higher pre-menopausal BMI was associated with a lower risk of ER-positive compared with ER-negative disease among White women, but this pattern was not observed in the other populations. Among Asian women, there was no evidence that the association with risk for premenopausal BMI varied by subtype. The published estimates in populations of Black or African American women were inconclusive (due to high variability in reported associations) and there were too few studies in Hispanic or Latina women to evaluate heterogeneity.

**Figure 4.**
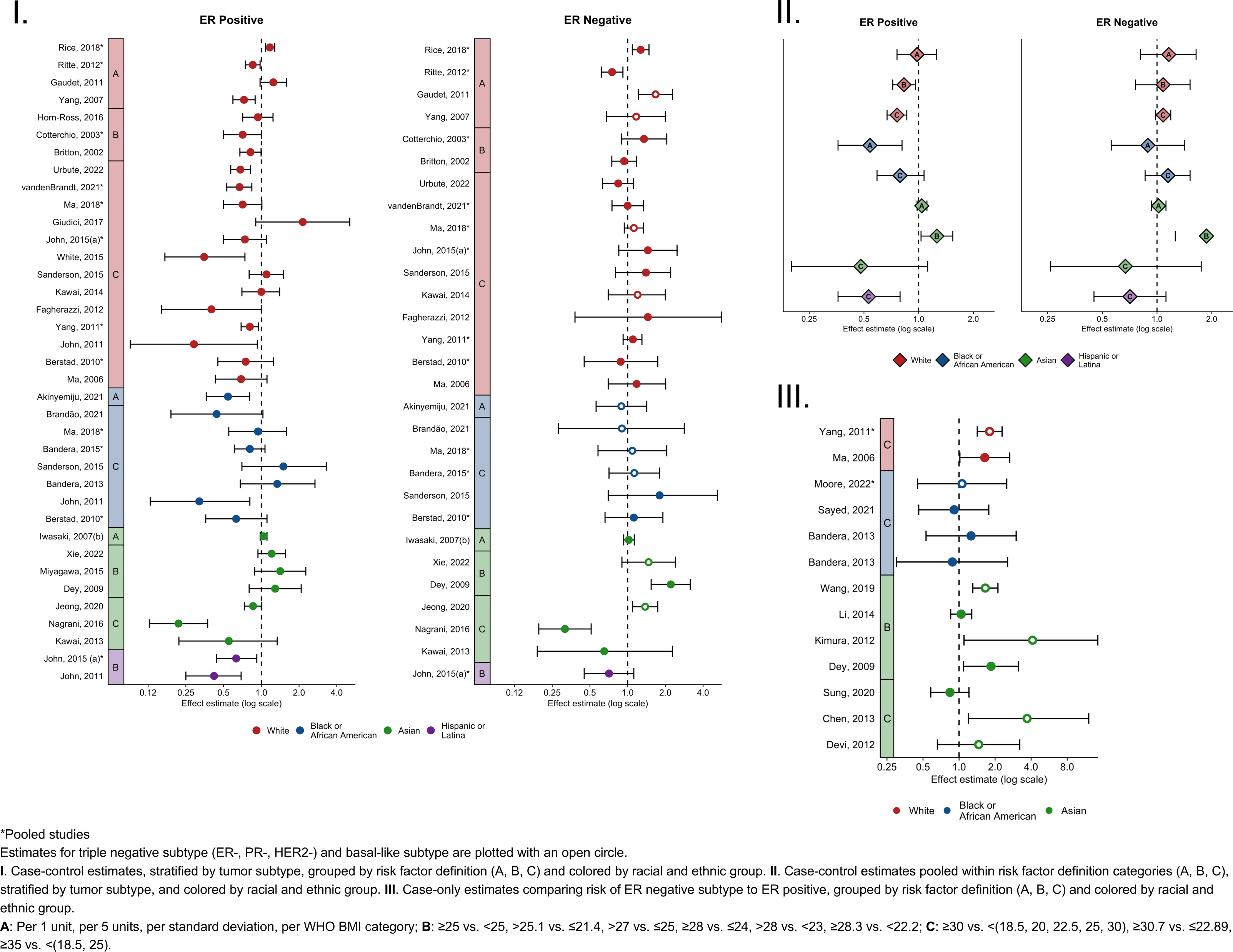
Published estimates for the effect of premenopausal BMI on breast cancer risk by tumor subtype and racial and ethnic group.

**Figure 5.**
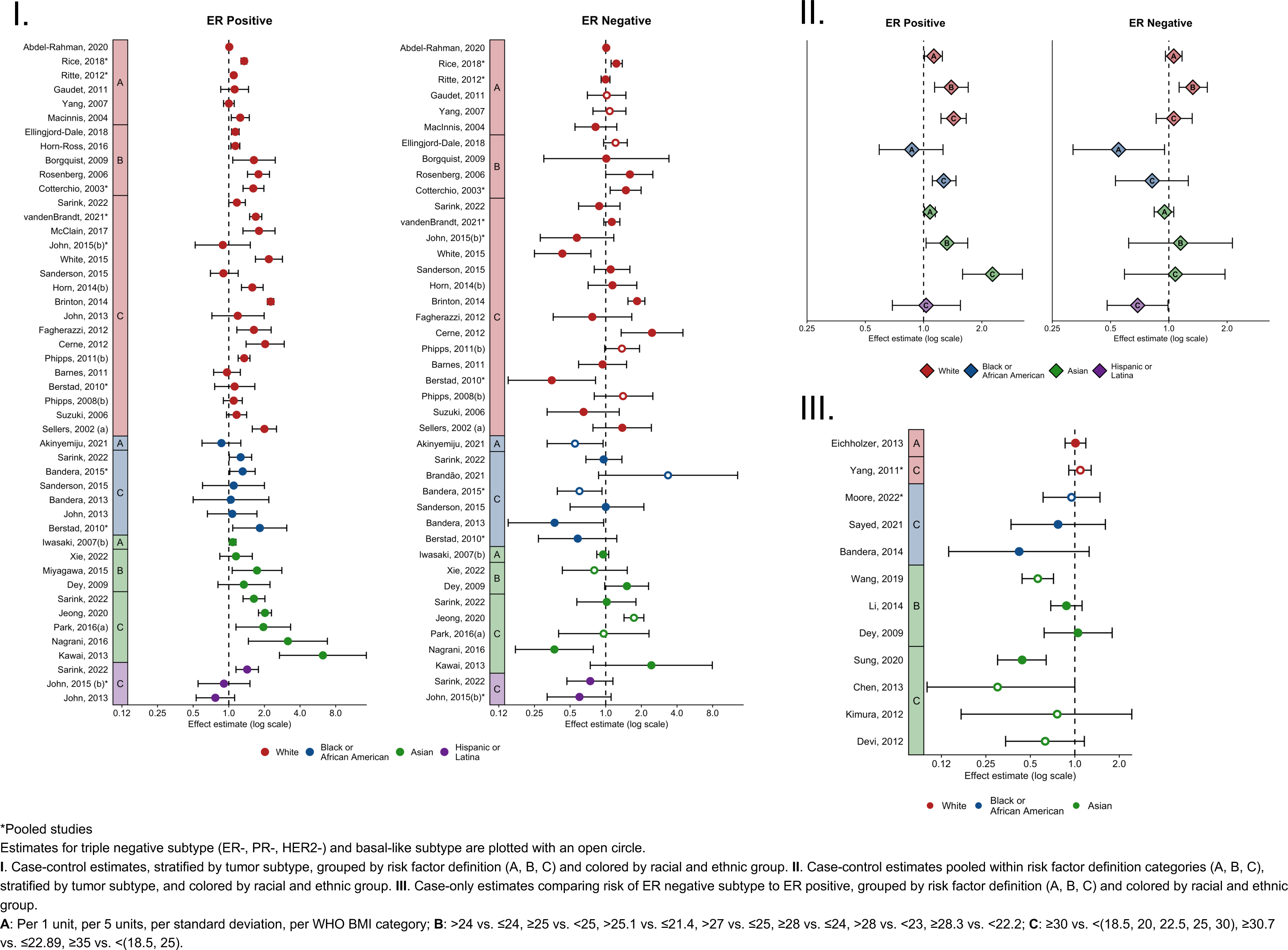
Published estimates for the effect of postmenopausal BMI on breast cancer risk by tumor subtype and racial and ethnic group.

### Lifestyle Factors

There were 37 eligible publications on alcohol intake, 30 on physical activity, and 25 on smoking status. There was no evidence of heterogeneity in risk by breast cancer subtype for any of these lifestyle risk factors. Increased alcohol use was associated with a higher risk of breast cancer, regardless of subtype, which was consistent across population groups (**Figure S8, Table S11**). History of smoking and higher physical activity were associated with a higher and lower risk of breast cancer, respectively, regardless of subtype (**Figures S9-S10, Tables S12-S13**). The evidence for smoking and physical activity was either inconclusive or not able to be assessed in populations of Asian or Latina descent due to too few published studies in these populations.

### Medical Factors/Medication Use

This review identified 41 publications reporting associations for first-degree family history of breast cancer, 30 on oral contraceptive use, 22 on mammographic breast density, 17 on use of combined (estrogen and progesterone) MHT, 16 on use of estrogen-only MHT, and 14 on history of benign breast disease. Few studies evaluating heterogeneity of medical factors/medication use were published in non-White populations, with particularly few studies among populations of Asian, or Hispanic or Latina descent.

Having a first-degree relative with a history of breast cancer was associated with a higher risk of ER-positive and -negative disease, which was consistent across race and ethnicity groups (**Figure S11, Table S14**). Similarly, there was no evidence of etiologic heterogeneity for risk associated with benign breast disease or mammographic density; however, almost all the published evidence on these factors was in White populations (**Figures S12-S13, Tables S15-S16**).

The evidence was suggestive of etiologic heterogeneity for use of oral contraceptives and menopausal hormone therapy among some racial and ethnic populations. Among White and Black or African American women, use of oral contraceptives was associated with an increased risk of ER-negative disease and minimal-to-no increased risk of ER-positive disease (**Figure S14**, **Table S17**). This pattern was most evident among studies of longer duration of use (≥10 years). Evidence of heterogeneity was inconclusive in populations of Asian and Hispanic or Latina descent due to relatively few published studies and high variability across studies.

The evidence was suggestive of etiologic heterogeneity for the use of estrogen-only MHT in White women, with an increased risk of ER-positive and no association with ER-negative disease (**Figure S15**, **Table S18**). Among Black or African American populations, however, estrogen-only MHT use was not associated with risk of either tumor subtype. This contrasts with combined MHT (estrogen and progesterone) use, for which the evidence was suggestive of heterogeneity among Black or African American populations (**Figure S16**, **Table S19**). Use of combined MHT was associated with an increased risk of breast cancer among White women, but the evidence for etiologic heterogeneity was inconclusive. There were too few studies of MHT use among Asian or Hispanic or Latina women to evaluate heterogeneity in risk by subtype in these groups.

## DISCUSSION

This review uncovered convincing to suggestive evidence to support heterogenous risk associations across ER subtypes for parity, and suggestive to inconclusive evidence for other established breast cancer risk factors (age at first birth, BMI, OC use, and MHT use). For other risk factors, the evidence of etiologic heterogeneity by ER-defined subtypes was either absent or inconclusive. This review also identified gaps in the literature that warrant further investigation. Findings were generally consistent across racial and ethnic groups, with the strongest evidence coming from studies including White populations.

### Etiologic heterogeneity across subtypes

Our findings are consistent with prior reviews and meta-analyses in that the associations between reproductive risk factors and risk of breast cancer subtypes show strong evidence for heterogeneity of risk associations by ER status. Reviews of parity^9,10,23–27^ and age at first birth^9–11,23–26^ generally report associations with ER-positive subtype, but no consistent relationships with ER-negative. The biological mechanisms explaining the relationship between parity, age at first birth, and risk of breast cancer subtypes are likely multifactorial. Increased risk of ER-positive disease among nulliparous women is thought to be due to “uninterrupted” exposure to endogenous sex hormones,^28,29^ and multiparity is thought to be protective due to its role in inducing the terminal differentiation of luminal epithelial cells, downregulation of growth factors, and upregulation of growth inhibitory signals.^30,31^ However, detailed studies of the association between time since last birth and breast cancer risk have found parity associated with a transient increase in risk right after birth that is attenuated with time.^32,33^

Heterogeneity in risk associations between breastfeeding and ER-defined subtypes has been inconsistent in prior reviews/meta-analyses, with most reporting similar inverse associations with both subtypes,^9,10,23–25^ while two recent reviews reported stronger inverse associations for triple negative compared to luminal breast cancers.^26,34^ The inverse association of breastfeeding with risk of ER-positive and ER-negative breast cancers suggests multiple mechanisms at play. Hormonal mechanisms (e.g., absence of ovulatory menstrual cycles and shorter lifetime exposure to endogenous sex hormones) may, in part, explain the reduced risk of ER-positive disease among women who have breastfed.^35^ Possible mechanisms for the inverse association of breastfeeding on risk of ER-negative disease include permanent changes to the breast histology, demonstrated by involution of terminal duct lobular units (TDLU); although, TDLU involution has also been associated with a lower risk of ER-positive disease ^36,37^

Consistent with the current review, prior reviews of age at menarche and subtype-specific breast cancer risk generally found similar inverse associations with both subtypes.^9,23,24,26,38^ There have been relatively few prior reviews on subtype-specific associations with age at menopause, and these have had inconsistent results. One reported a stronger association for ER-positive compared to ER-negative cases,^38^ with a more recent meta-analysis reporting an association only among luminal cases (with no difference between subtypes in case-case analyses).^26^ Similar to this recent meta-analysis, we did not find sufficient evidence to conclude a difference in association for age at menopause across subtypes.

Prior reviews and meta-analyses of the associations between pre- and postmenopausal BMI with risk of breast cancer subtypes are limited, but are generally consistent with the findings of the current review. Prior reviews reported that higher BMI is inversely associated with ER-positive/PR-positive disease among premenopausal women, but associated with a higher risk of ER-positive/PR-positive disease among postmenopausal women.^14,23,39^ Associations between pre- and post-menopausal BMI and ER-negative subtypes have been unclear or null. Differential associations of high postmenopausal BMI and risk of breast cancer subtypes is biologically intuitive, as after menopause estrogen production takes place in the adipose tissue, creating a favorable microenvironment for the development of ER-positive tumors.^40,41^ The reason for the opposite pattern—high premenopausal BMI being associated with reduced risk of ER-positive subtype—is not yet clear. A possible contributing factor is that chronic inflammation influences breast physiology differently depending on menopausal status, implicating a role for estrogens and adipokine-driven signaling pathways.^42^ Additionally, with estrogen synthesis prior to menopause occurring mainly in the ovaries, the higher prevalence of irregular/anovulatory cycles among obese women can lead to decreased circulating hormone levels and decreased risk of ER-positive disease.^42–45^

Prior reviews and meta-analyses on subtype-specific associations with oral contraceptive use have generally agreed that use is associated with an increased risk of ER-negative breast cancer.^23,26,46,47^ Two prior reports conclude that the evidence is unclear.^24,25^ These same reviews have found either no association, or a potential inverse association between OC use and risk of ER-positive breast cancer. The biological mechanism explaining the heterogeneity in association by tumor subtype is not completely clear. One hypothesis is that estrogen promotes the growth of ER-negative breast cancers by systematically increasing angiogenesis and stromal cell recruitment.^48^

The previous reviews and meta-analyses have also consistently shown that use of either estrogen-only or combined menopausal hormone therapy is associated with increased risk of ER-positive breast cancer.^15,49,50^ Associations with ER-negative breast cancer have been less consistently reported. While the present review found suggestive evidence of subtype heterogeneity among some populations, this is based on published evidence of current/former/never use. There is evidence that risk increases with longer duration of use, and the risk is greater for ER-positive than for ER-negative for any given duration of use.

### Etiologic heterogeneity across racial and ethnic groups

Although the distributions of breast cancer risk factors are known to vary across groups, there is currently little or no evidence that relative risk estimates for established risk factors by ER-subtypes differ substantially across racial or ethnic groups. However, there is a need for larger and comparable studies to rule out differences in relative risk particularly for studies in non-White populations and of non-reproductive risk factors. Studies among Hispanic and Latina women are particularly limited.

### Limitations of the review

Our review has several limitations, which largely reflect the constraints in the current body of published literature on breast cancer subtype etiologic heterogeneity. The inconsistencies in how risk factors were categorized across publications, as well as in how the breast cancer subtypes were defined, make comparisons difficult. This variation in exposure and outcome definitions across publications prevented us from performing a meta-analysis across published estimates, and therefore our conclusions are based on a consensus review, rather than statistical testing. Also, there is limited literature on subtypes defined by other markers or characteristics (e.g., tumor grade, *TP53*); thus, we only reviewed publications that defined tumor subtypes based on ER status or “surrogate” subtypes. Some reports demonstrate that there are additional sources of etiologic heterogeneity that are not well captured by ER status.^51–54^

While we included most established breast cancer risk factors in this review, there are certain factors that we could not report due to a lack of published studies (particularly in non-White populations). These include the cross-classification of parity and breastfeeding, time since last birth, time from menarche to first birth, type of benign breast disease, duration of MHT use, modification of the effect of BMI by MHT use, and others. Studies evaluating the role of body composition measures by menopausal status and by race and ethnicity are also lacking.

We recognize that the racial and ethnic groups included in this review represent a simplification of diverse populations which represent many languages, countries, continents, and cultures and do not address complex social variables that may represent differential exposure patterns across groups. For example, the Asian group included publications representing women from East Asia (e.g., China, Japan), South Asia (e.g., India), Southeast Asia (e.g., Malaysia), West Asia (e.g., Turkey, Iraq), and self-identifying Asian American women. Additionally, indigenous African populations (e.g., from Ghana, Mozambique, Rwanda, and Kenya) were categorized as Black, Latin American populations (e.g., from Brazil, Mexico, Chile, and Colombia) were categorized as Hispanic or Latino; however, most studies of Black or African American women, as well as Hispanic or Latina women included US populations that self-identify into these groups. The publications of White women also encompass a diverse group, including studies from the US, UK, Germany, Slovenia, Australia, Canada, etc. Efforts should be made by future studies to attempt to disaggregate these highly heterogenous racial and ethnic groups.

Scoping reviews are warranted when the literature on a particular topic has not yet been comprehensively reviewed or when the body of literature is not suited for a more precise systematic review due to its large or heterogeneous nature.^55,56^ Assessment of study quality is not considered mandatory for a scoping review and completing a formal bias assessment would be very difficult in this case; moreover, assessing bias was not a focus of our review. We acknowledge, however, that not all publications are of equal quality. When we see outliers in the published literature, this may be due to bias, and it highlights the importance of including many studies to get the clearest picture of the observed patterns.

### Conclusions and recommendations for future work

Uncovering the sources of breast cancer etiologic heterogeneity is of great importance to improve disease prevention and early detection, particularly for tumor subtypes associated with low survival, which remain major contributors to breast cancer mortality globally. The field has focused mainly on defining breast cancer subtypes using ER status or combining multiple markers (ER, PR, HER2) to form molecular subtypes when evaluating etiologic heterogeneity. While there are clear differences in treatment strategies between ER-positive and ER-negative breast tumors, the optimal, most etiologically relevant schema for defining breast cancer subtypes may not necessarily be rooted in treatment differences. For instance, studies have shown associations of individual risk factors with other molecular markers, such as histologic grade, KI67, and TP53 mutation status within luminal breast cancers.^51–54^ Accordingly, a change in approach may be warranted to better characterize etiologic subtypes of breast cancer with epidemiologic and public health relevance.

Future work to evaluate etiologic heterogeneity across breast cancer subtypes will require large, high-quality epidemiological studies in diverse populations with comprehensive and standardized data on risk factors and pathology subtypes, as well as greater adoption of FAIR (findable, accessible, interoperable, and reusable) data principles to improve reproducibility of findings and to facilitate data pooling across studies.^57–59^ Larger, high-quality epidemiological studies with detailed data on risk factors and tumor characteristics are needed to refine the relative risk estimates in specific tumor subtypes, and in specific populations—particularly in populations underrepresented in current studies. Additional studies evaluating possible heterogeneity in subtype-specific risks across racial and ethnic groups are needed, especially those that account for social variables that may modify risk and that differ by race or ethnicity. This need is particularly noteworthy for non-reproductive risk factors.

## Supporting information

Supplemental References

## CONFLICTS OF INTEREST

The authors have no conflicts of interest to disclose.

## FUNDING

This work was supported by Intramural Funds of the National Cancer Institute, USA. MGC is supported by Breast Cancer Now and the Institute of Cancer Research, UK. RK and MKS were supported by the European Union’s Horizon 2020 Research and Innovation Program B-CAST (grant number: 633784). BDJ and ARH are supported by the Cancer Prevention Fellowship Program.

## DATA AVAILABILITY

The datasets were derived from sources in the public domain. All data are incorporated into the article and its online supplementary material.

## Notes

### Competing Interest Statement

The authors have declared no competing interest.

