## Supplementary figures and images for "Risk factors for breast cancer subtypes by race and ethnicity: A scoping review of the literature"

### Figure S1

**Supplementary Figure 1**. Screening flow chart

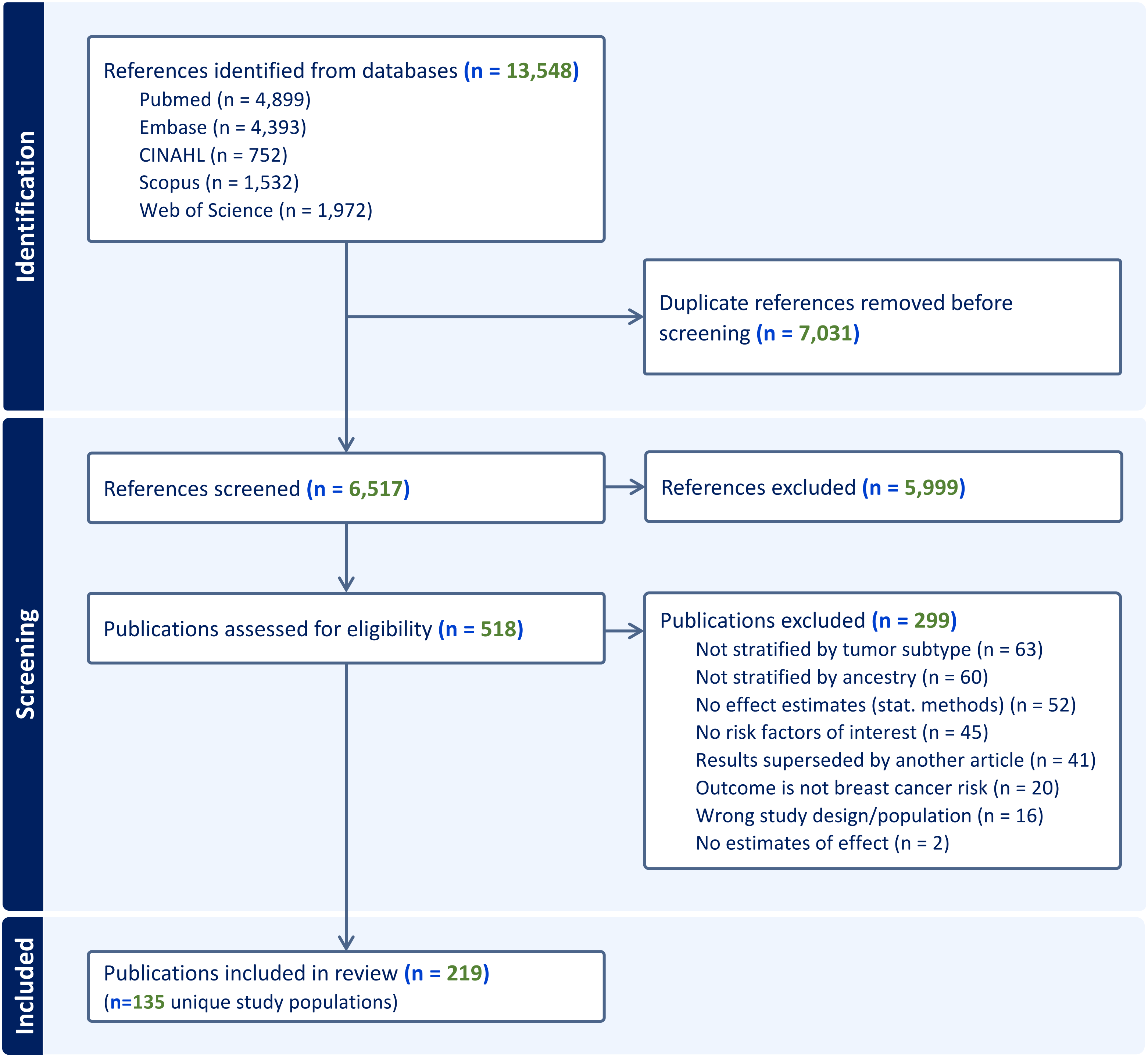

### Figure S2

**Supplementary Figure 2**. Criteria for determining evidence of subtype heterogeneity

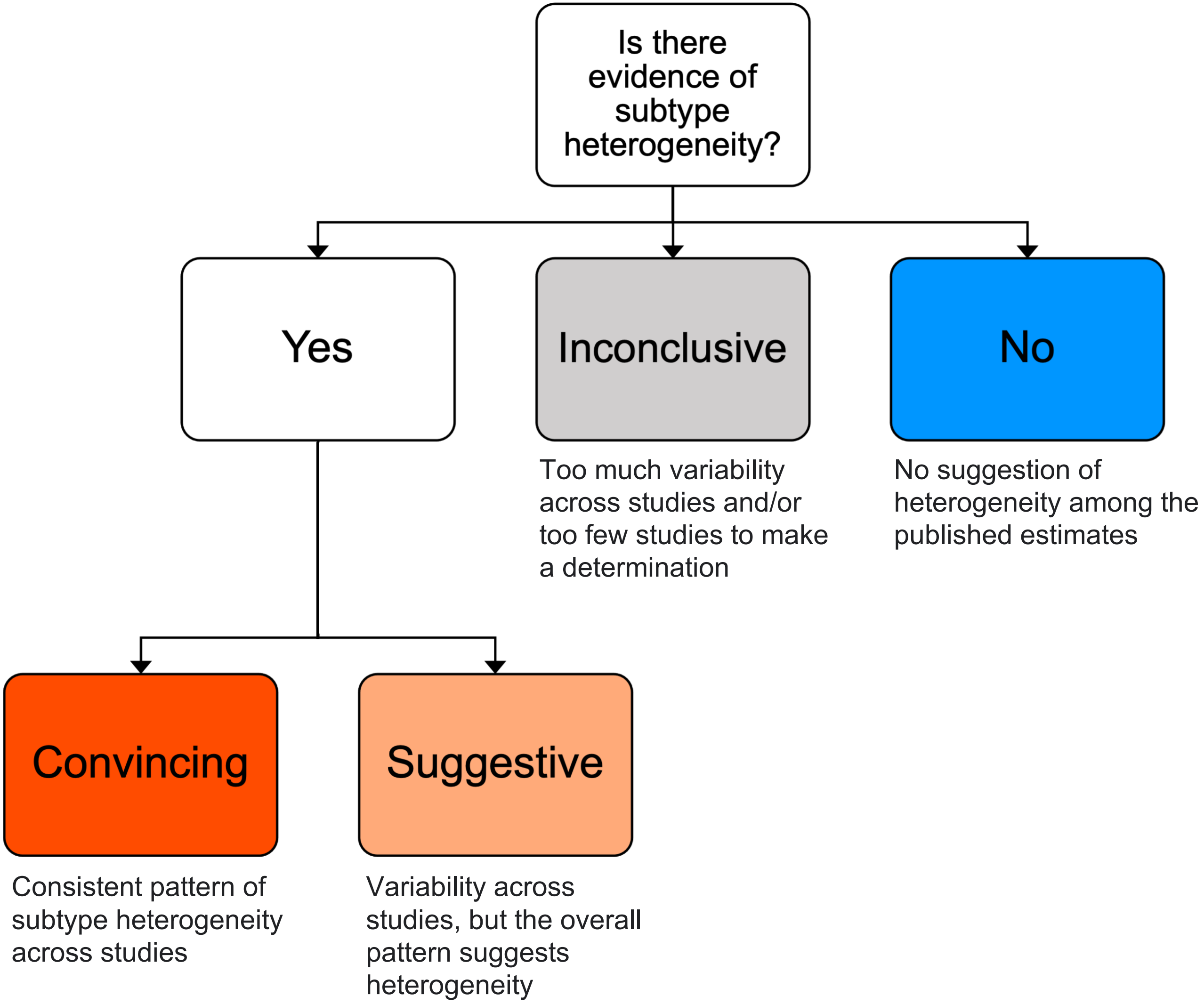
