## Supplementary material for "Risk factors for breast cancer subtypes by race and ethnicity: A scoping review of the literature": Figure S3

**Supplementary Figure 3**. Published estimates for the effect of **age at menarche** on breast cancer risk by tumor subtype and racial and ethnic group

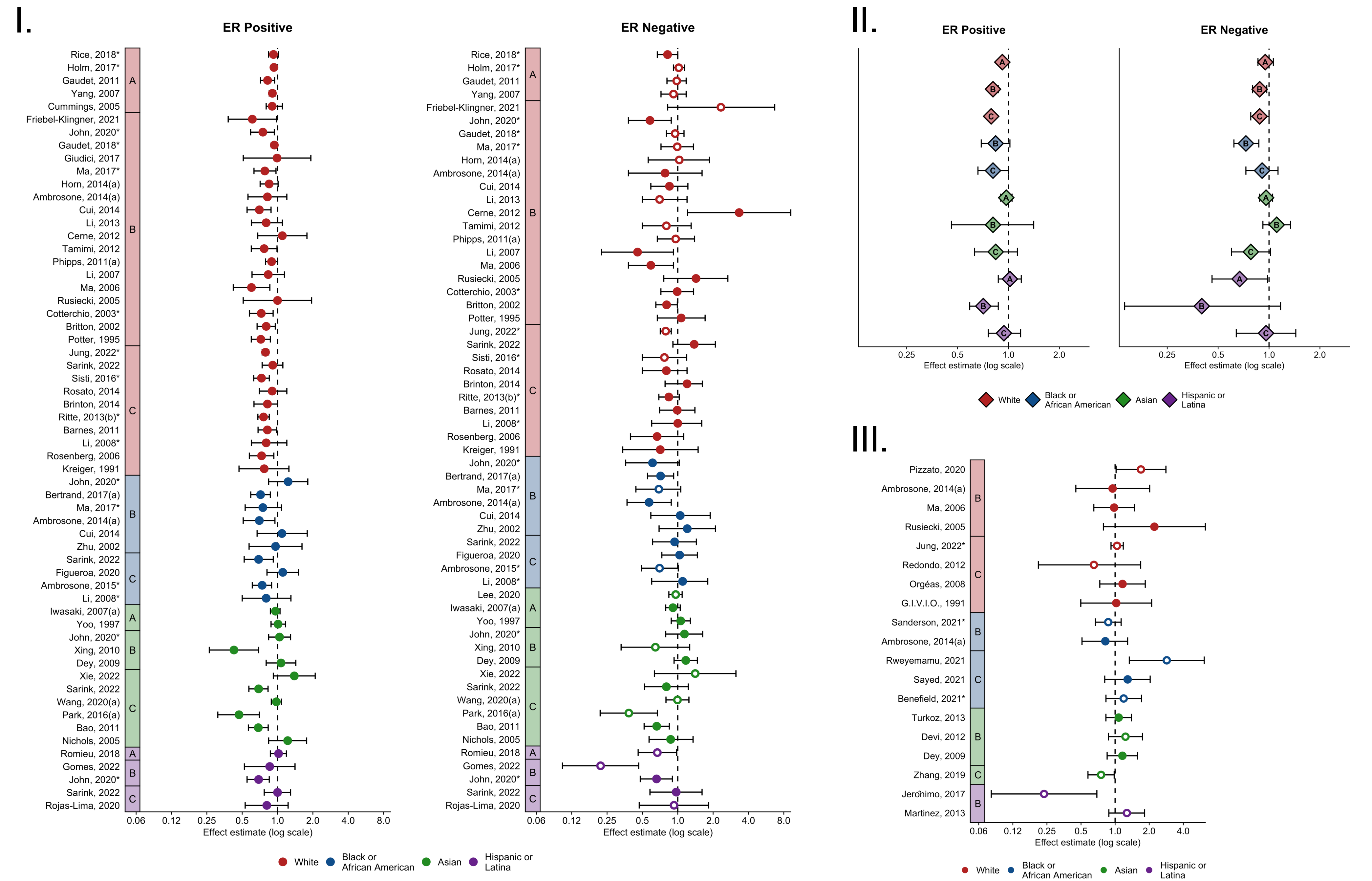


*Pooled studies

Estimates for triple negative subtype (ER-, PR-, HER2-) and basal-like subtype are plotted with an open circle.

**I**. Case-control estimates, stratified by tumor subtype, grouped by risk factor definition (A, B, C) and colored by racial and ethnic group. **II**. Case-control estimates pooled within risk factor definition categories (A, B, C), stratified by tumor subtype, and colored by racial and ethnic group. **III**. Case-only estimates comparing risk of ER negative subtype to ER positive, grouped by risk factor definition (A, B, C) and colored by racial and ethnic group.

**A**: Per 1 year, per 1.5 years, per 2 years; **B**: ≥12 vs. <12 years, ≥13 vs. <13 years, ≥14 vs. <(12, 13, 14) years; **C**: ≥15 vs. <(11, 12, 13, 14) years, ≥16 vs. <(12, 13, 14) years, ≥17 vs. <(14, 15) years, ≥18 vs. <(14, 15) years.
