## Supplementary material for "Risk factors for breast cancer subtypes by race and ethnicity: A scoping review of the literature": Figure S4

**Supplementary Figure 4**. Published estimates for the effect of **breastfeeding** on breast cancer risk by tumor subtype and racial and ethnic group

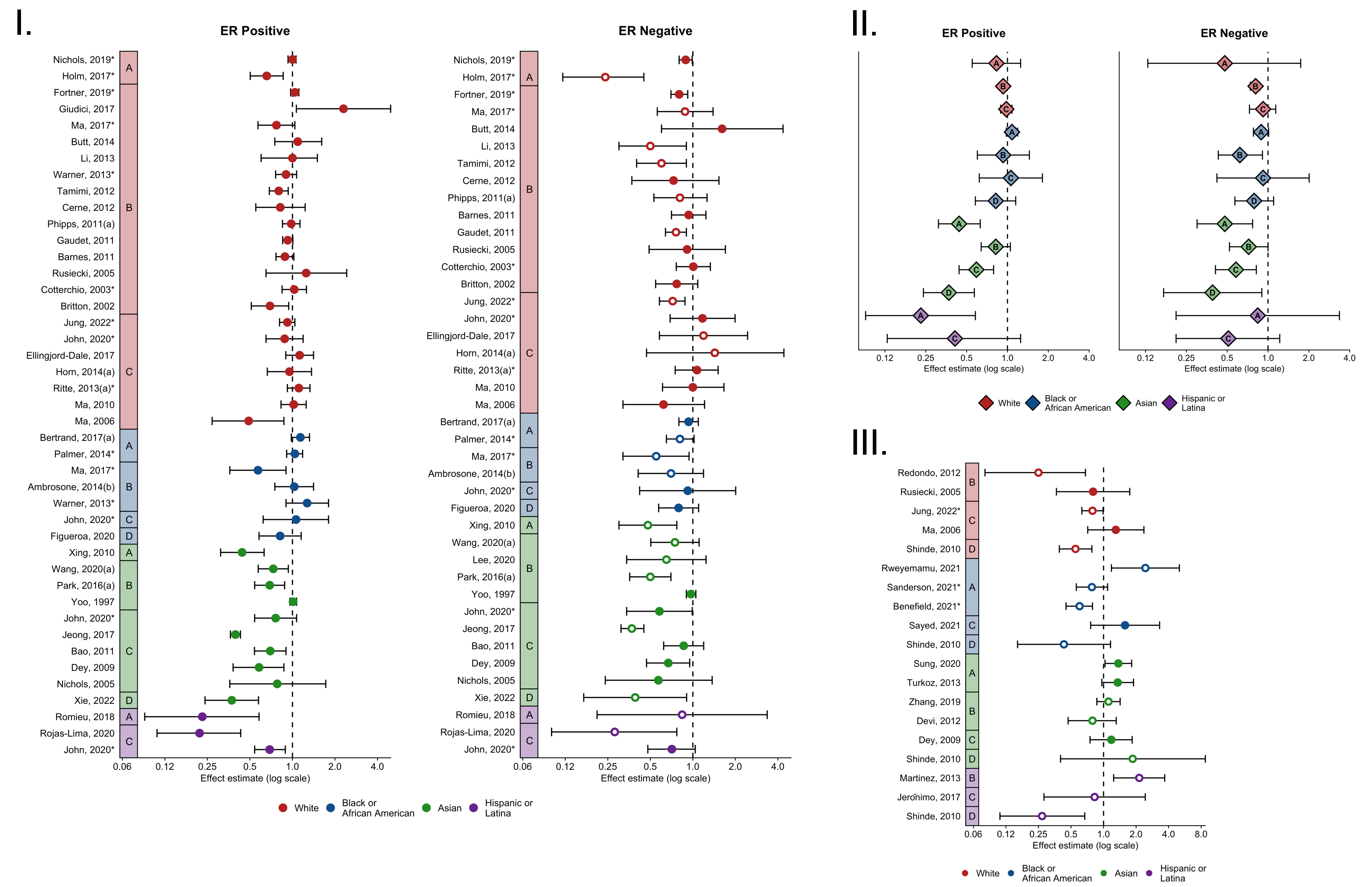


*Pooled studies

Estimates for triple negative subtype (ER-, PR-, HER2-) and basal-like subtype are plotted with an open circle.

**I**. Case-control estimates, stratified by tumor subtype, grouped by risk factor definition (A, B, C, D) and colored by racial and ethnic group. **II**. Case-control estimates pooled within risk factor definition categories (A, B, C, D), stratified by tumor subtype, and colored by racial and ethnic group. **III**. Case-only estimates comparing risk of ER negative subtype to ER positive, grouped by risk factor definition (A, B, C, D) and colored by racial and ethnic group.

**A**: Ever vs. Never; **B**: Per 3 months, per 6 months, ≥4 vs. 0 months, ≥6 vs. 0 months, ≥7 vs. (0,1-6) months, ≥12 vs. 0 months, ≥13 vs. Nulliparous, ≥13 vs. (0, <4) months; **C**:≥18 vs. (0, <1) months, ≥24 vs. 0 months, ≥25 vs. (0, 1-3) months, ≥31 vs. 0 months, ≥61 vs. <12 months, ≥73 months vs. Nulliparous, ≥79 vs. <36 months; **D:** ≥19 vs.<13 median months/pregnancy, <7 months/birth.
