## Supplementary material for "Risk factors for breast cancer subtypes by race and ethnicity: A scoping review of the literature": Figure S5

**Supplementary Figure 5**. Published estimates for the effect of **age at menopause** on breast cancer risk by tumor subtype and racial and ethnic group

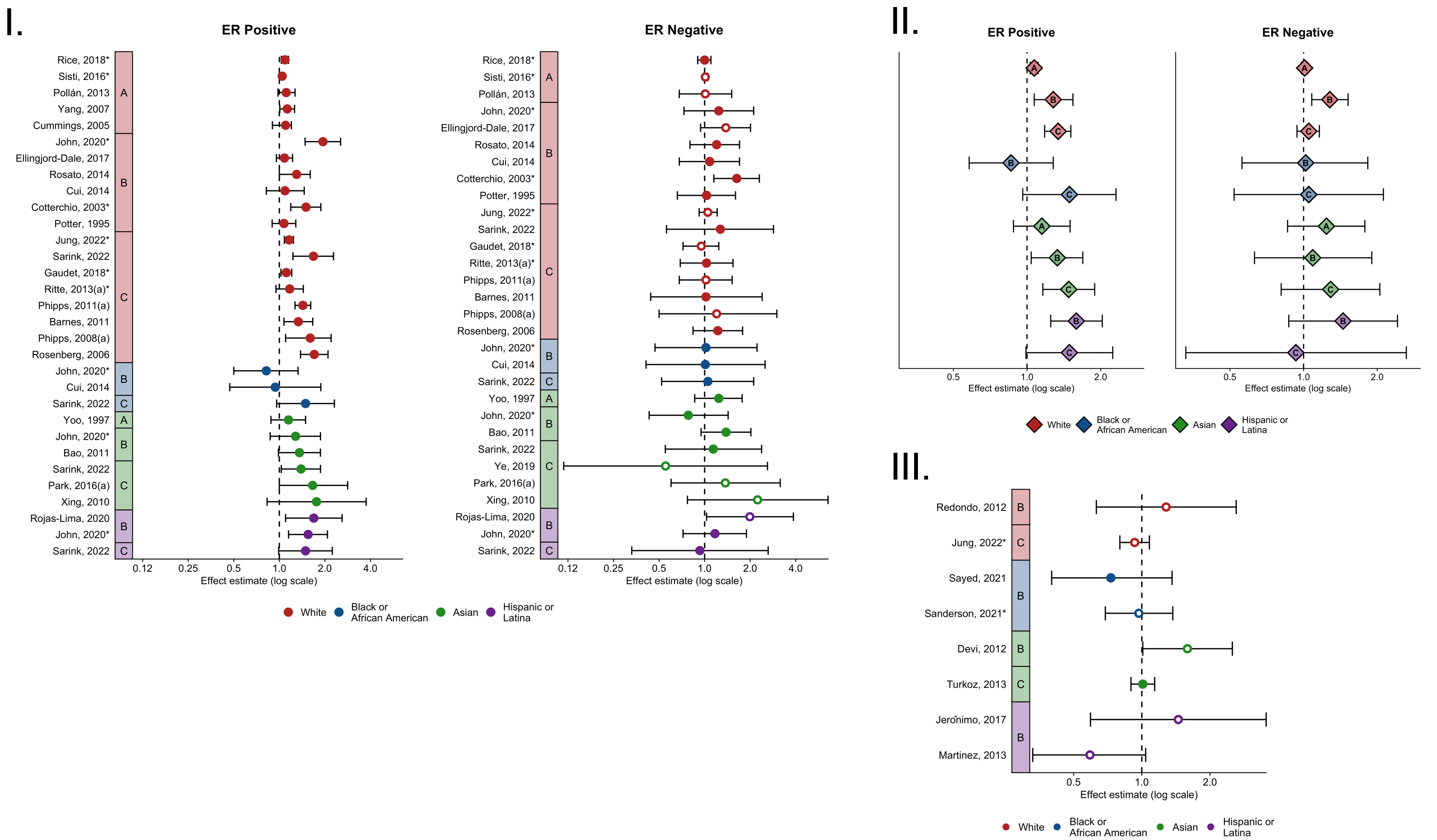


*Pooled studies

Estimates for triple negative subtype (ER-, PR-, HER2-) and basal-like subtype are plotted with an open circle.

**I**. Case-control estimates, stratified by tumor subtype, grouped by risk factor definition (A, B, C, D) and colored by racial and ethnic group. **II**. Case-control estimates pooled within risk factor definition categories (A, B, C, D), stratified by tumor subtype, and colored by racial and ethnic group. **III**. Case-only estimates comparing risk of ER negative subtype to ER positive, grouped by risk factor definition (A, B, C, D) and colored by racial and ethnic group.

**A**: Per 1 year, per 5 years, per 6.3 years, per category increase; **B**: ≥49 vs. <43 years, ≥50 vs. <(45, 50) years, ≥51 vs. <(46, 51) years, ≥52 vs. <(40, 47) years, ≥53 vs. <47 years; **C**: ≥54 vs. <50 years, ≥55 vs. <(35, 44, 45, 49, 50, 55) years.
