## Supplementary material for "Risk factors for breast cancer subtypes by race and ethnicity: A scoping review of the literature": Figure S6

**Supplementary Figure 6**. Published estimates for the effect of **height** on breast cancer risk by tumor subtype and racial and ethnic group

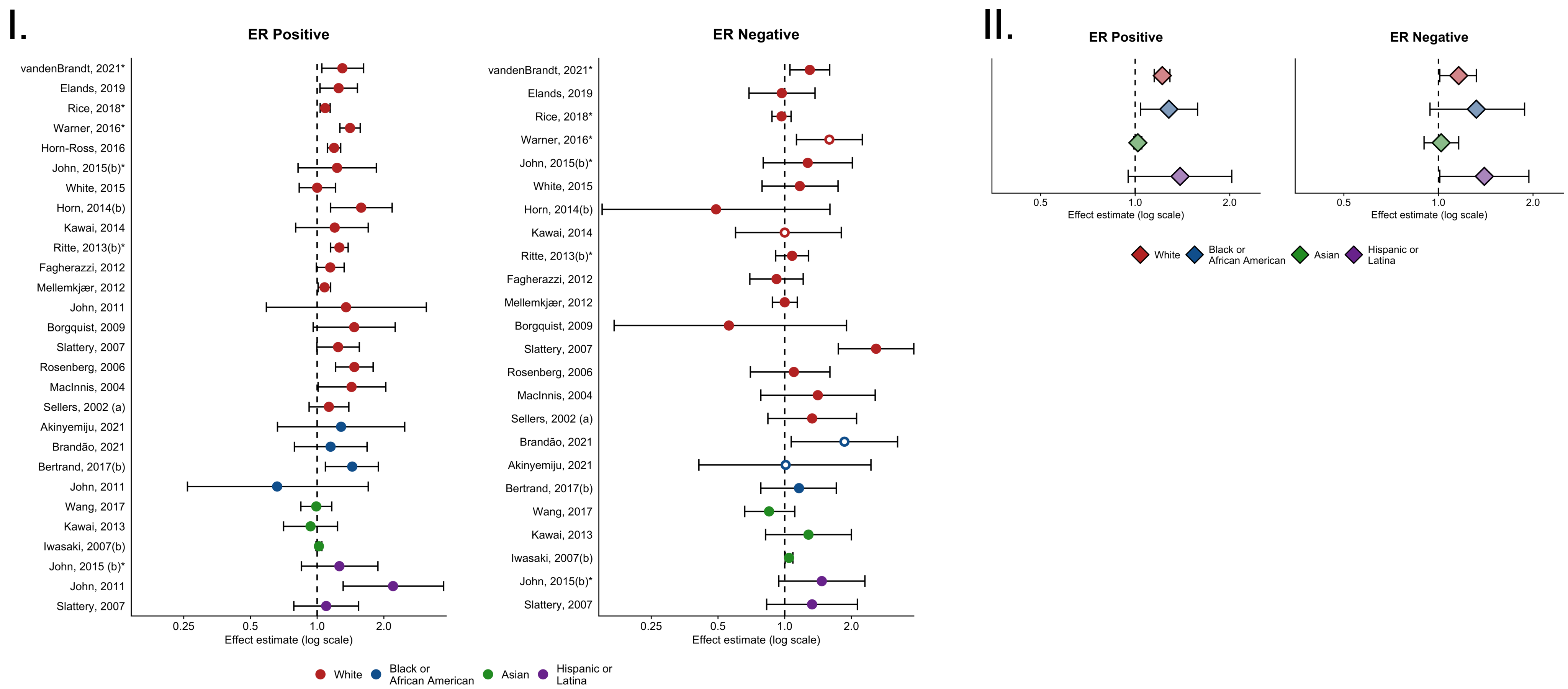


*Pooled studies

Estimates for triple negative subtype (ER-, PR-, HER2-) and basal-like subtype are plotted with an open circle.

**I.** Case-control estimates, stratified by tumor subtype colored by racial and ethnic group. **II**. Case-control estimates pooled within and colored by racial and ethnic group, stratified by tumor subtype.

Estimates defined as per (5 cm, 10 cm, 3 in), ≥157 vs. <149 cm, >159.4 vs. <151.2 cm, >160 vs. ≤155 cm, >160.2 vs. ≤160.2 cm, ≥164 vs. <159 cm, >164 vs. ≤156 cm, >166.6 vs. ≤161.8 cm, >168 vs. <160 cm, ≥170 vs. <160 cm,
