## Supplementary material for "Risk factors for breast cancer subtypes by race and ethnicity: A scoping review of the literature": Figure S7

**Supplementary Figure 7**. Published estimates for the effect of **waist-to-hip ratio** on breast cancer risk by tumor subtype and racial and ethnic group

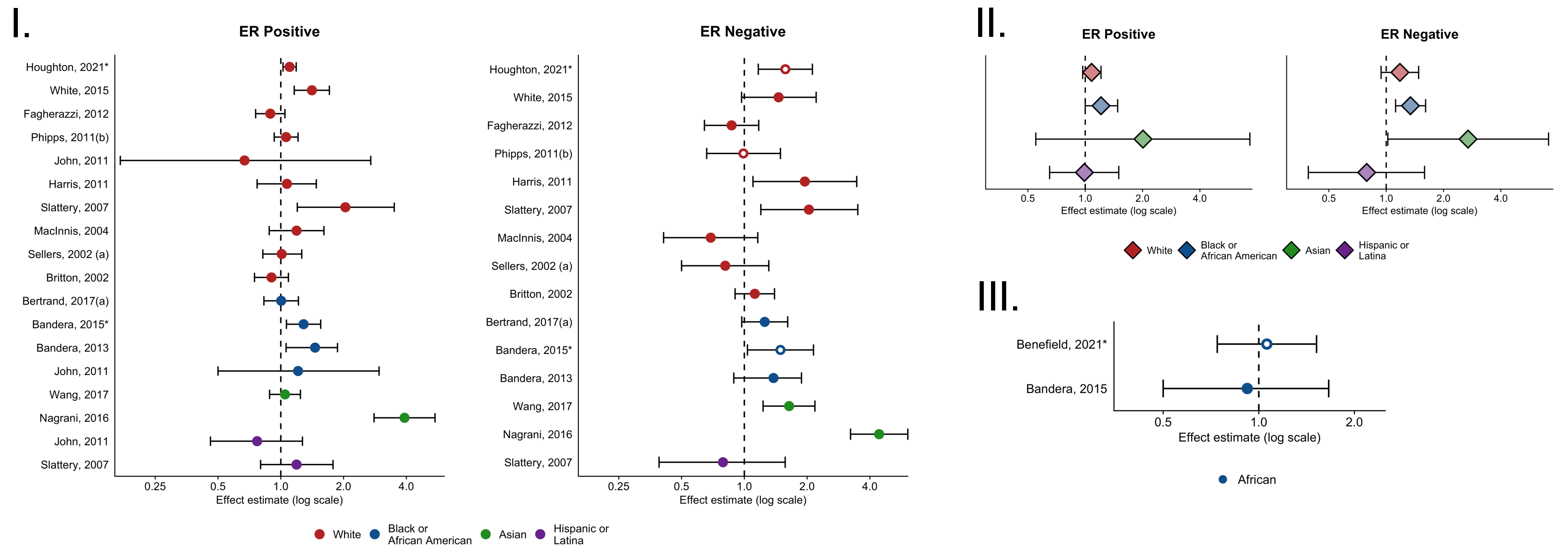


*Pooled studies

Estimates for triple negative subtype (ER-, PR-, HER2-) and basal-like subtype are plotted with an open circle.

**I.** Case-control estimates, stratified by tumor subtype colored by racial and ethnic group. **II**. Case-control estimates pooled within and colored by racial and ethnic group, stratified by tumor subtype. **III**. Case-only estimates comparing risk of ER negative subtype to ER positive, colored by racial and ethnic group.

Estimates correspond to Per 0.1 units, ≥0.80 vs. <0.75, ≥0.84 vs. <0.73, ≥0.85 vs. <(0.76, 0.80), ≥0.86 vs. <0.86, ≥0.87 vs. <0.758, ≥0.89 vs. <0.75, ≥0.91 vs. <(0.77, 0.80), ≥0.93 vs. <(0.83, 0.93), ≥0.95 vs. <0.85, 90th vs. 10th percentile.
