## Supplementary material for "Risk factors for breast cancer subtypes by race and ethnicity: A scoping review of the literature": Figure S8

**Supplementary Figure 8**. Published estimates for the effect of **alcohol intake** on breast cancer risk by tumor subtype and racial and ethnic group

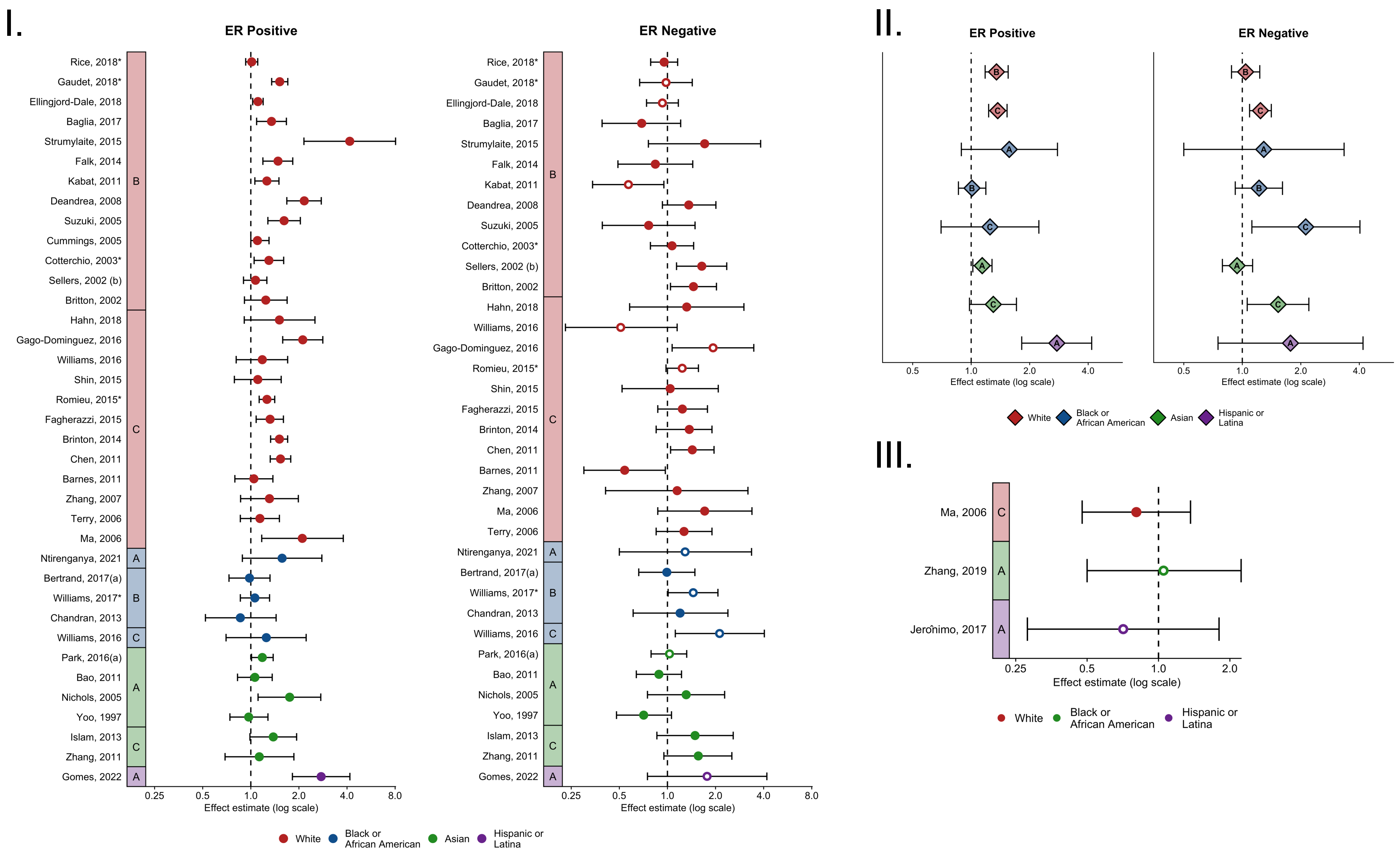


*Pooled studies

Estimates for triple negative subtype (ER-, PR-, HER2-) and basal-like subtype are plotted with an open circle.

**I**. Case-control estimates, stratified by tumor subtype, grouped by risk factor definition (A, B, C) and colored by racial and ethnic group. **II**. Case-control estimates pooled within risk factor definition categories (A, B, C), stratified by tumor subtype, and colored by racial and ethnic group. **III**. Case-only estimates comparing risk of ER negative subtype to ER positive, grouped by risk factor definition (A, B, C) and colored by racial and ethnic group.

**A**: Ever vs. Never, Any vs. None, Yes vs. No; **B**: ≥3 vs. 0 drinks/day, ≥3 vs. <3 drinks/week, ≥3.5 vs. 0 drinks/week, per 4 drinks/week, ≥6 vs. 0 drinks/week, ≥7 vs. (0, <0.5, <1) drinks/week, ≥5 vs. 0 g/day, per 10 g/day, ≥10 vs. 0 g/day, ≥13.8 g/day vs. Never, ≥28 vs. <14 g/week; **C**: ≥8 drinks/week vs. Never, ≥8 vs. 0 drinks/week, ≥13 vs. 0 drinks/week, ≥15 vs. 0 g/day, ≥23 vs. 0 g/day, ≥30 vs. 0 g/day, >30 vs. 0 g/day, >35 vs. 0 g/day.
