## Supplementary material for "Risk factors for breast cancer subtypes by race and ethnicity: A scoping review of the literature": Figure S9

**Supplementary Figure 9**. Published estimates for the effect of **smoking** on breast cancer risk by tumor subtype and racial and ethnic group

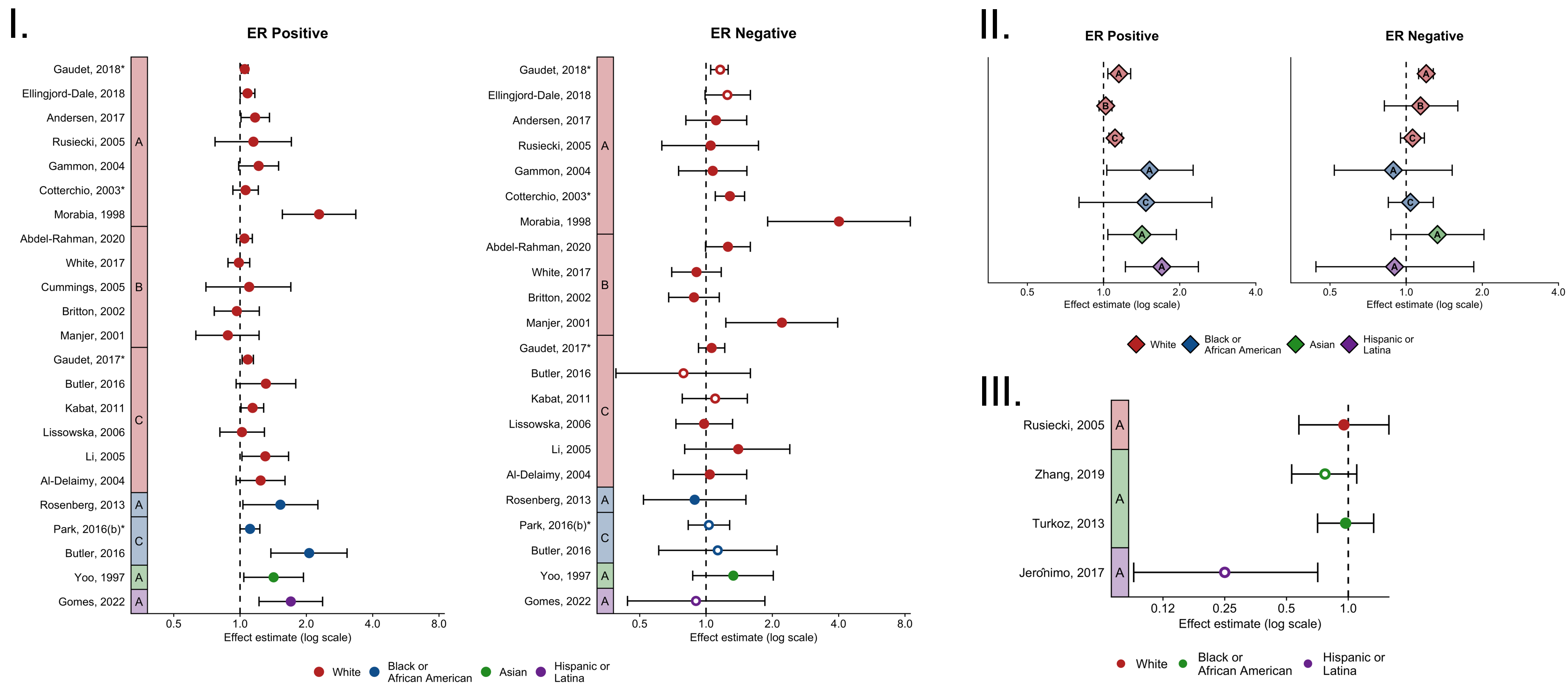


*Pooled studies

Estimates for triple negative subtype (ER-, PR-, HER2-) and basal-like subtype are plotted with an open circle.

**I**. Case-control estimates, stratified by tumor subtype, grouped by risk factor definition (A, B, C) and colored by racial and ethnic group. **II**. Case-control estimates pooled within risk factor definition categories (A, B, C), stratified by tumor subtype, and colored by racial and ethnic group. **III**. Case-only estimates comparing risk of ER negative subtype to ER positive, grouped by risk factor definition (A, B, C) and colored by racial and ethnic group.

**A**: Ever vs. Never; **B**: Current vs. Never, Current vs. non-current; **C**: ≥(20,21) vs. 0 years, ≥21 vs. 0 years, ≥30 vs. 0 years, ≥40 vs. 0 years.
