## Supplementary material for "Risk factors for breast cancer subtypes by race and ethnicity: A scoping review of the literature": Figure S10

**Supplementary Figure 10**. Published estimates for the effect of **physical activity** on breast cancer risk by tumor subtype and racial and ethnic group

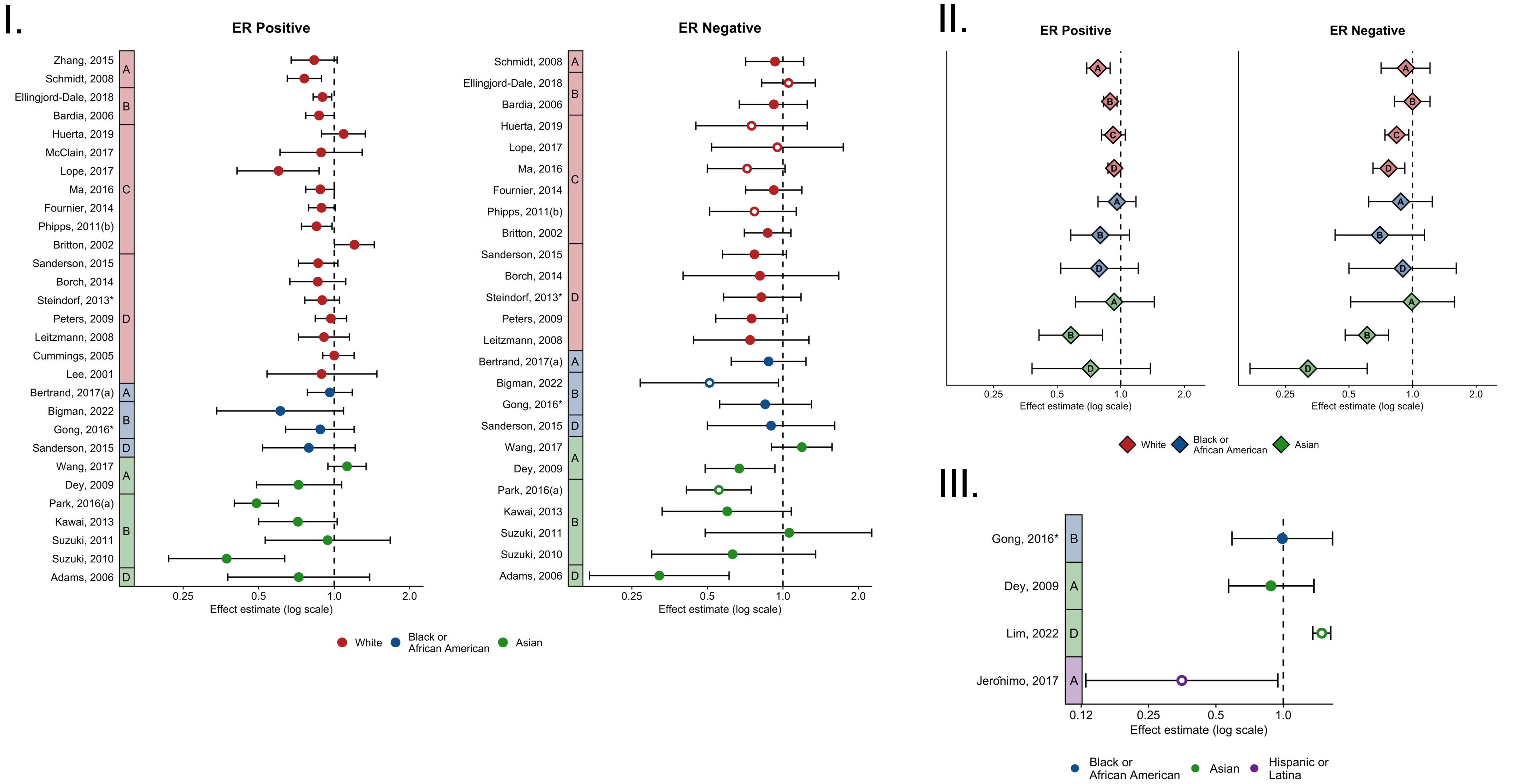


*Pooled studies

Estimates for triple negative subtype (ER-, PR-, HER2-) and basal-like subtype are plotted with an open circle.

**I**. Case-control estimates, stratified by tumor subtype, grouped by risk factor definition (A, B, C, D) and colored by racial and ethnic group. **II**. Case-control estimates pooled within risk factor definition categories (A, B, C, D), stratified by tumor subtype, and colored by racial and ethnic group. **III**. Case-only estimates comparing risk of ER negative subtype to ER positive, grouped by risk factor definition (A, B, C, D) and colored by racial and ethnic group.

**A: Leisure-time physical activity** (≥1 vs. 0 days/week, ≥1 vs. 0 days/week of moderate intensity, ≥2 times/week of vigorous intensity of ≥5 times/week of moderate intensity vs. <1 times/week of moderate intensity, ≥3 days/week vs. ≤3 days/month, ≥4 vs. <4 hours/week, ≥5 vs. <1 hours/week, ≥14.75 vs. <3.75 MET-hours/week); **B: Recreational physical activity** (>7 vs. ≤0.5 hours/week/year, ≥13.46 vs. 0 hours/week, ≥12 vs. <12 MET-hours/week, ≥16.5 vs. 0 MET-hours/week, >36 vs. ≤12 MET-hours/week, >1,000 vs. 0 MET-hours/week, >13.5 vs. ≤13.5 relative units/week); **C: Total physical activity** (Per 1,631 kcal/week, ≥4,200 vs. 0 kJ/week, active vs. inactive, very high vs. very low, active ≥5 times/week vs. inactive, any duration of vigorous intensity vs. ≤2 hours/week of light intensity, >0.87 vs. 0 MET-hours/day/year, ≥2.3 vs. 0 MET-hours/day, ≥395 vs. <244 MET-hours/week); **D: Other/type not specified** (Yes vs. no, ≥5 vs. <5 hours/week of brisk walking, >6 vs. <3 hours/day of physical activity, ≥5 vs. 0 hours/week of vigorous exercise).
