## Supplementary material for "Risk factors for breast cancer subtypes by race and ethnicity: A scoping review of the literature": Figure S11

**Supplementary Figure 11**. Published estimates for the effect of **family history of breast cancer** on breast cancer risk by tumor subtype and racial and ethnic group

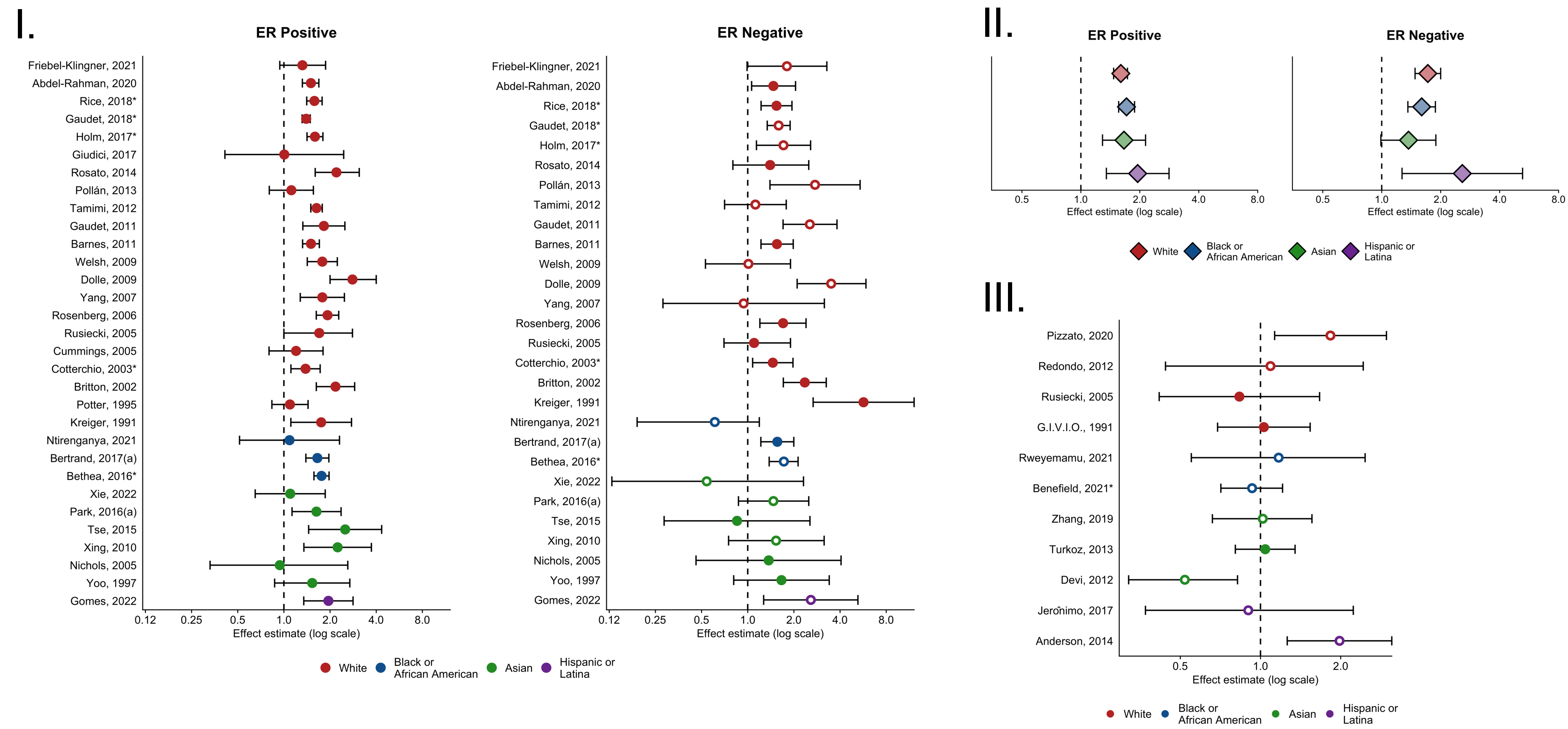


*Pooled studies

Estimates for triple negative subtype (ER-, PR-, HER2-) and basal-like subtype are plotted with an open circle.

**I.** Case-control estimates, stratified by tumor subtype colored by racial and ethnic group. **II**. Case-control estimates pooled within and colored by racial and ethnic group, stratified by tumor subtype. **III**. Case-only estimates comparing risk of ER negative subtype to ER positive, colored by racial and ethnic group.

All estimates correspond to yes vs. no family history of breast cancer in first degree relatives.
