## Supplementary material for "Risk factors for breast cancer subtypes by race and ethnicity: A scoping review of the literature": Figure S12

**Supplementary Figure 12**. Published estimates for the effect of **history of benign breast disease** on breast cancer risk by tumor subtype and racial and ethnic group

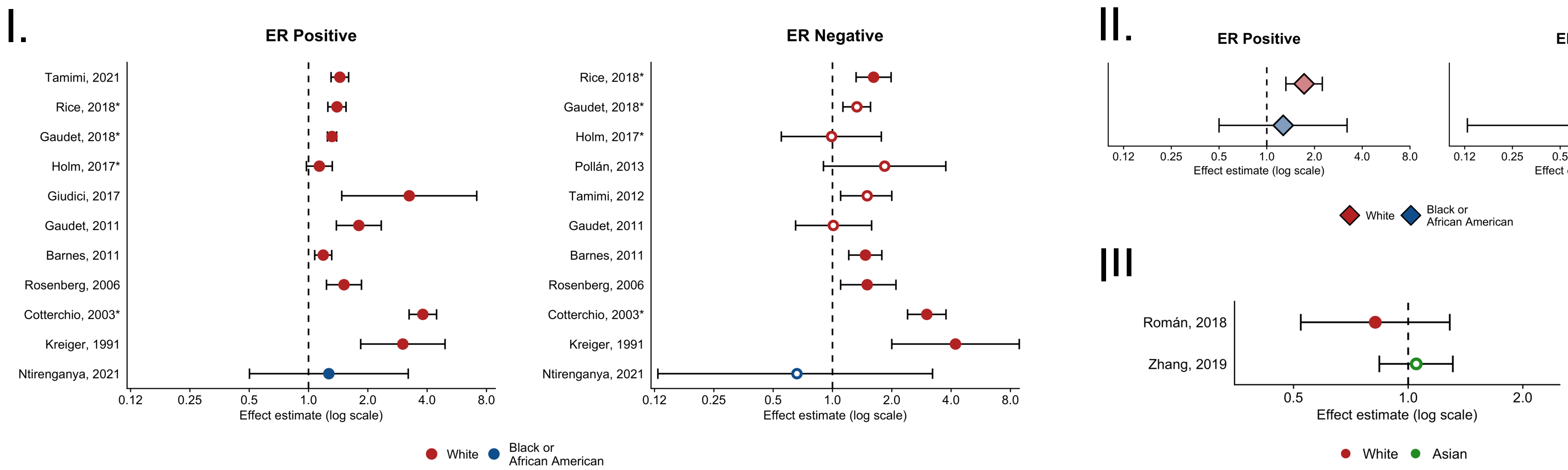


*Pooled studies

Estimates for triple negative subtype (ER-, PR-, HER2-) and basal-like subtype are plotted with an open circle.

**I.** Case-control estimates, stratified by tumor subtype colored by racial and ethnic group. **II**. Case-control estimates pooled within and colored by racial and ethnic group, stratified by tumor subtype. **III**. Case-only estimates comparing risk of ER negative subtype to ER positive, colored by racial and ethnic group.

Estimates correspond to yes vs. no and previous breast biopsy vs. none.
