## Supplementary material for "Risk factors for breast cancer subtypes by race and ethnicity: A scoping review of the literature": Figure S13

**Supplementary Figure 13**. Published estimates for the effect of **mammographic density** on breast cancer risk by tumor subtype and racial and ethnic group

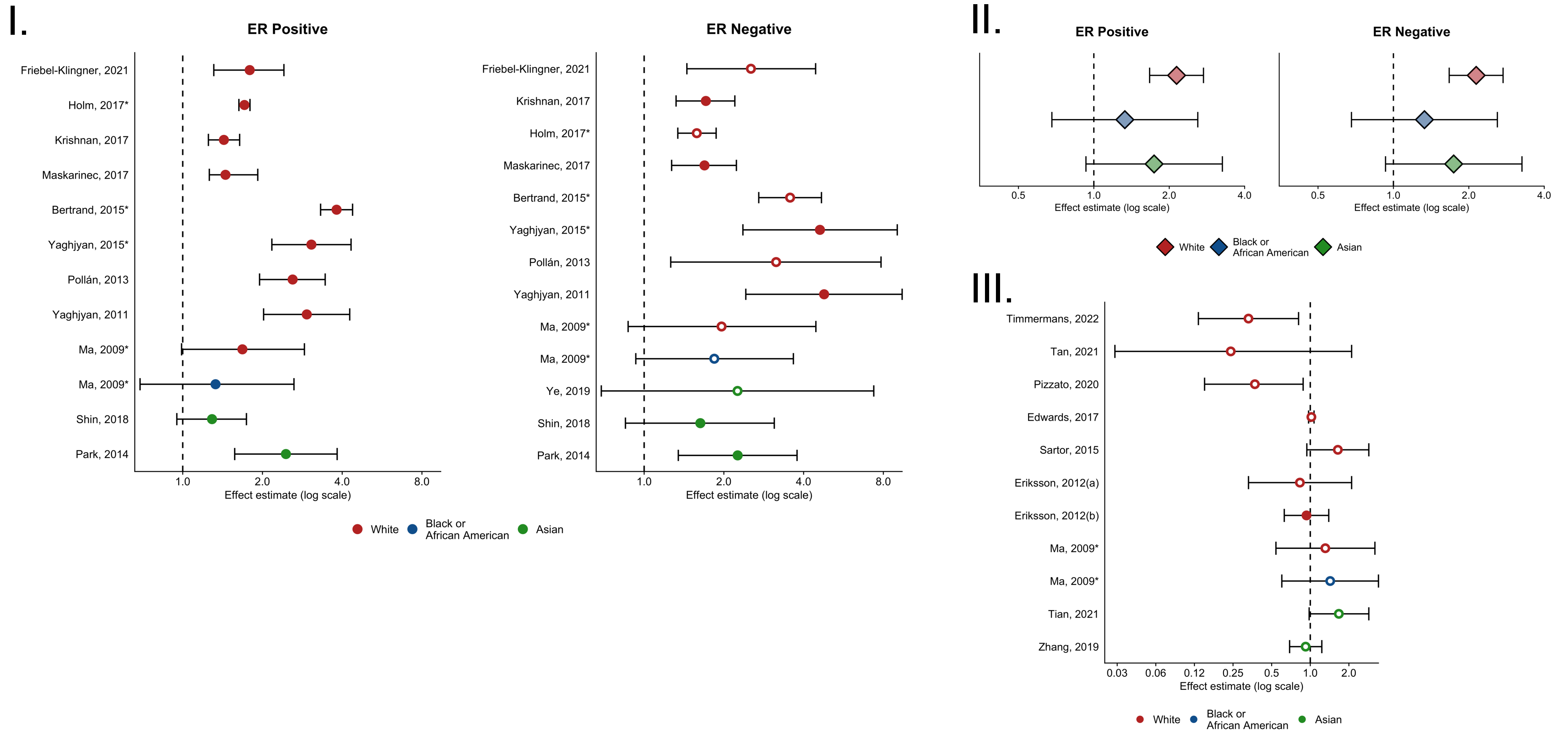


*Pooled studies

Estimates for triple negative subtype (ER-, PR-, HER2-) and basal-like subtype are plotted with an open circle.

**I.** Case-control estimates, stratified by tumor subtype colored by racial and ethnic group. **II**. Case-control estimates pooled within and colored by racial and ethnic group, stratified by tumor subtype. **III**. Case-only estimates comparing risk of ER negative subtype to ER positive, colored by racial and ethnic group.

Estimates defined as BIRADS dense vs. non-dense, BIRADS 4 vs. 1, BIRADS 3&4 vs. 1, BIRADS extremely vs. less than extremely dense, BIRADS heterogeneously/extremely dense vs. almost entirely fatty/scattered, Per BIRADS category, ≥50% vs. <10% density, >50% vs. 25-11% density, ≥30% vs. <30% density, ≥26.27 vs. <9% density, per SD of percent dense area, per SD of absolute dense area, per 1% volumetric density, VDG 4 vs. 1&2, VDG D vs. A&B.
