## Supplementary material for "Risk factors for breast cancer subtypes by race and ethnicity: A scoping review of the literature": Figure S15

**Supplementary Figure 15**. Published estimates for the effect of **estrogen-only menopausal hormone therapy use** on breast cancer risk by tumor subtype and racial and ethnic group

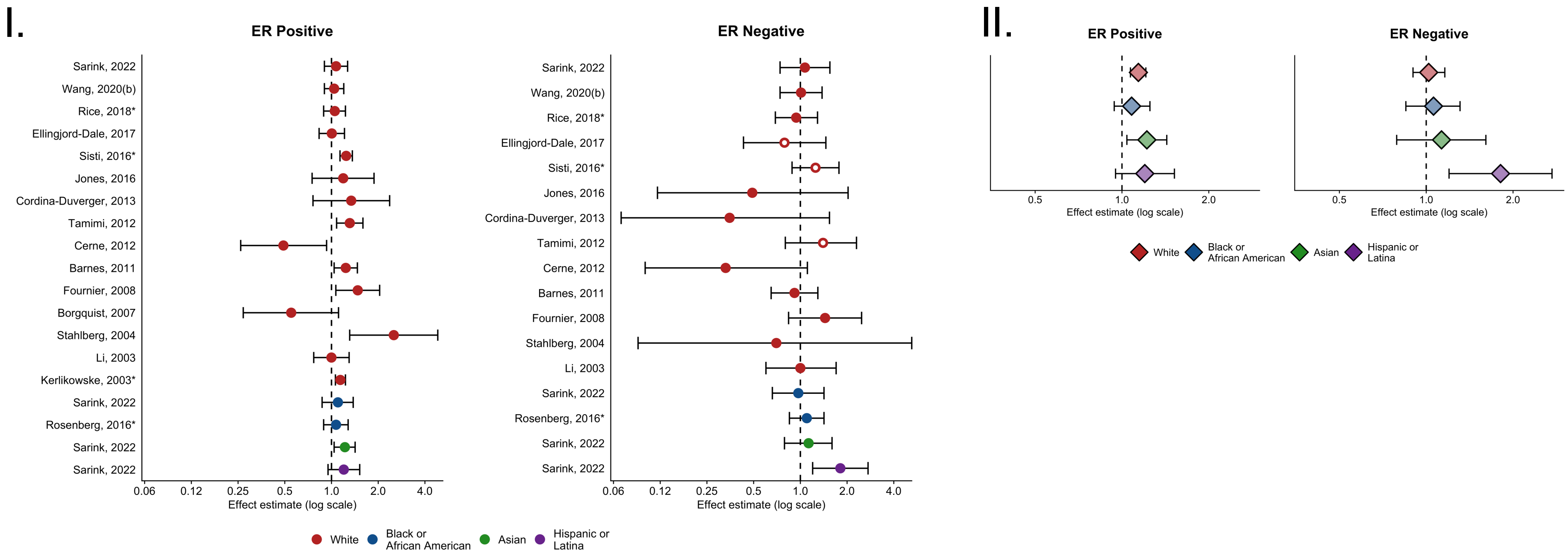


*Pooled studies

Estimates for triple negative subtype (ER-, PR-, HER2-) and basal-like subtype are plotted with an open circle.

**I.** Case-control estimates, stratified by tumor subtype colored by racial and ethnic group. **II**. Case-control estimates pooled within and colored by racial and ethnic group, stratified by tumor subtype.

Estimates correspond to ever vs. never use, current vs. never use, and current vs. non-current use.
