## Supplementary material for "Risk factors for breast cancer subtypes by race and ethnicity: A scoping review of the literature": Figure S16

**Supplementary Figure 16**. Published estimates for the effect of **combined menopausal hormone therapy use** on breast cancer risk by tumor subtype and racial and ethnic group

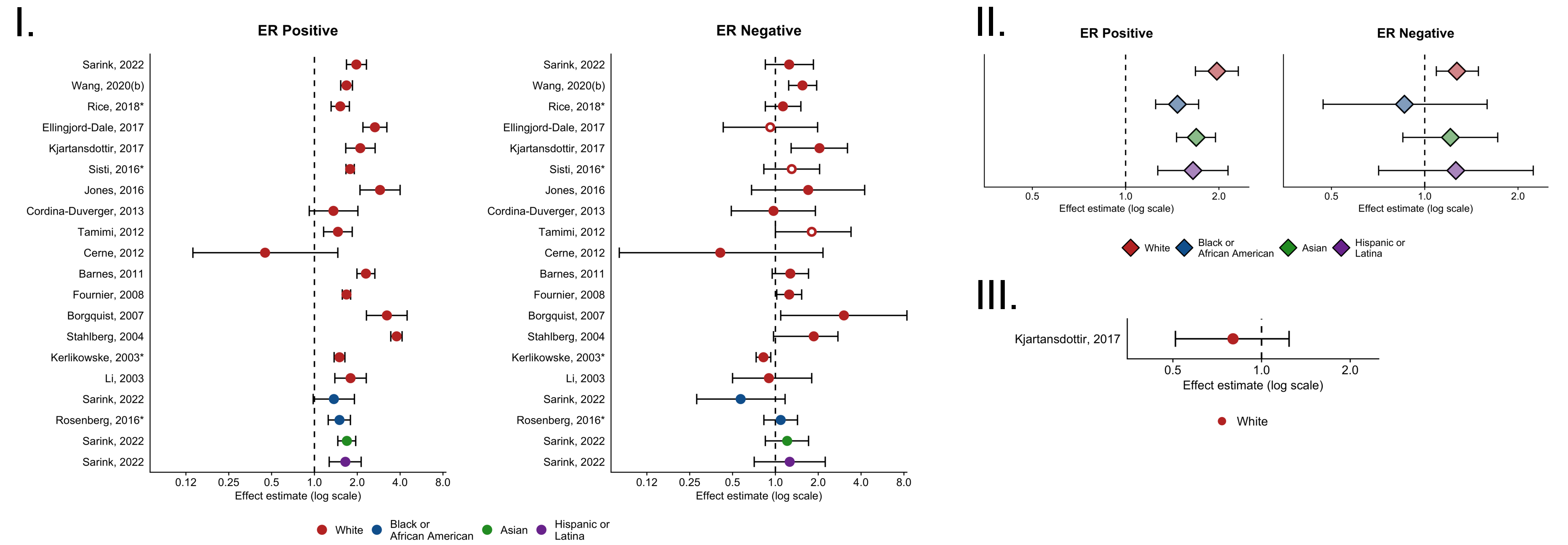


*Pooled studies

Estimates for triple negative subtype (ER-, PR-, HER2-) and basal-like subtype are plotted with an open circle.

**I.** Case-control estimates, stratified by tumor subtype colored by racial and ethnic group. **II**. Case-control estimates pooled within and colored by racial and ethnic group, stratified by tumor subtype. **III**. Case-only estimates comparing risk of ER negative subtype to ER positive, colored by racial and ethnic group.

Estimates correspond to ever vs. never use, current vs. never use, and current vs. non-current use.
