## Supplementary material for "Risk factors for breast cancer subtypes by race and ethnicity: A scoping review of the literature": Table S1

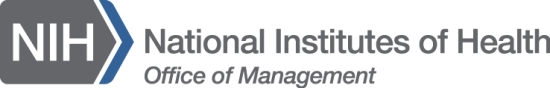

Database Search Strategy

Database: PubMed/MEDLINE

Platform: National Library of Medicine
Date Searched: 7/1/2022
Date: 1/1/1990-7/1/2022

Limit: English | Human/exclude animal studies and non-peer-reviewed

|  | Concept: | Search Strategy: |
| --- | --- | --- |
| #1 | Breast Cancer | "Breast Neoplasms"[MeSH Terms] OR "breast neoplasm*"[Title/Abstract] OR "breast cancer*"[Title/Abstract] |
| #2 | Exposures | "Menarche"[MeSH Terms] OR "menarche*"[Title/Abstract] OR "Menstruation"[MeSH Terms] OR "menstrua*"[Title/Abstract] OR "age at first birth"[Title/Abstract] OR "age at first childbirth"[Title/Abstract] OR "parity"[Title/Abstract] OR "parous"[Title/Abstract] OR "Breast Feeding"[MeSH Terms] OR "breastfeed*"[Title/Abstract] OR "Lactation"[MeSH Terms] OR "Lactation"[Title/Abstract] OR "lactating"[Title/Abstract] OR "oral contraceptive*"[Title/Abstract] OR "Hormone Replacement Therapy"[MeSH Terms] OR "hormone therap*"[Title/Abstract] OR "hormone replacement*"[Title/Abstract] OR "estrogen replacement*"[Title/Abstract] OR "progesterone therap*"[Title/Abstract] OR "progesterone replacement*"[Title/Abstract] OR "postmenopausal hormon*"[Title/Abstract] OR "age at menopause"[Title/Abstract] OR "menopausal age"[Title/Abstract] OR "menopausal status"[Title/Abstract] OR "Body Height"[MeSH Terms] OR "height"[Title/Abstract] OR "Body Mass Index"[MeSH Terms] OR "Body Mass Index"[Title/Abstract] OR "quetelet index"[Title/Abstract] OR "BMI"[Title/Abstract] OR "waist hip ratio"[MeSH Terms] OR "waist-to-hip"[Title/Abstract] OR "waist hip ratio"[Title/Abstract] OR "physical activit*"[Title/Abstract] OR "Alcohol Drinking"[MeSH Terms] OR "alcohol"[Title/Abstract] OR "Tobacco Smoking"[MeSH Terms] OR "smoking"[Title/Abstract] OR "family history"[Title/Abstract] OR "benign breast disease"[Title/Abstract] OR "BBD"[Title/Abstract] OR "mammographic densit*"[Title/Abstract] OR "breast densit*"[Title/Abstract] |
| #3 | Subtype | "subtype*"[Title/Abstract] OR "hormone receptor*"[Title/Abstract] OR "receptors, estrogen"[MeSH Terms] OR "estrogen receptor*"[Title/Abstract] OR "oestrogen receptor*"[Title/Abstract] OR "triple negative*"[Title/Abstract] OR "luminal*"[Title/Abstract] |
| #4 |  | #1 AND #2 AND #3 |
| #5 | Limits & Filters | (((#1 AND #2 AND #3) NOT ("Animals"[MeSH Terms] AND "Humans"[MeSH Terms]))) NOT ("mice"[Title/Abstract] OR "mouse"[Title/Abstract] OR "murine"[Title/Abstract] OR "rat"[Title/Abstract] OR "rats"[Title/Abstract] OR "rodent*"[Title/Abstract] OR "Rodentia"[MeSH Terms] OR "dog"[Title/Abstract] OR "Dogs"[Title/Abstract] OR "Dogs"[MeSH Terms] OR "pig"[Title/Abstract] OR "pigs"[Title/Abstract] OR "piglet*"[Title/Abstract] OR "swine*"[Title/Abstract] OR "Swine"[MeSH Terms] OR "porcine*"[Title/Abstract] OR "Animal Experimentation"[MeSH Terms] OR "models, animal"[MeSH Terms] OR "In Vitro Techniques"[MeSH Terms] OR "cell culture*"[Title/Abstract] OR "MCF7"[Title/Abstract] OR "T47D"[Title/Abstract] OR "MDAMB231"[Title/Abstract])) NOT ("letter"[Publication Type] OR "editorial"[Publication Type] OR "editorial"[Title/Abstract] OR "comment"[Publication Type] OR "news"[Publication Type] OR "Congress"[Publication Type] OR "Consensus Development Conference"[Publication Type] OR "conference abstract*"[Title/Abstract] OR "conference paper*"[Title/Abstract] OR "conference review*"[Title/Abstract] OR "conference proceeding*"[Title/Abstract] OR "retracted publication"[Publication Type] OR "retraction of publication"[Publication Type] OR "retraction of publication"[Title/Abstract] OR "retraction notice"[Title] OR "retracted publication"[Title/Abstract] OR "Published Erratum"[Publication Type] OR "corrigenda"[Title/Abstract] OR "corrigendum"[Title/Abstract] OR "errata"[Title/Abstract] OR "erratum"[Title/Abstract] OR "protocol"[Title] OR "protocols"[Title] OR "Case Reports"[Publication Type] OR "case report"[Title] OR "case series"[Title] OR "case stud*"[Title/Abstract])) AND ((1990/1/1:2022/7/1[pdat]) AND (english[Filter])) |

Database: Embase

Platform: Elsevier
Date Searched: 7/1/2022
Date: 1/1/1990-7/1/2022

Limit: English | Human/exclude animal studies and non-peer-reviewed

|  | Concept: | Search Strategy: |
| --- | --- | --- |
| #1 | Breast Cancer | 'breast cancer'/exp OR 'or breast neoplasm*':ab,ti OR 'breast cancer*':ab,ti |
| #2 | Exposures | 'menarche'/exp OR 'menstruation'/exp OR 'breast feeding'/exp OR 'lactation'/exp OR 'hormone substitution'/exp OR 'body height'/exp OR 'body mass'/exp OR 'waist hip ratio'/exp OR 'drinking behavior'/exp OR 'smoking'/exp OR 'menarche*':ab,ti OR 'menstrua*':ab,ti OR 'age at first birth':ab,ti OR 'age at first childbirth':ab,ti OR 'parity':ab,ti OR 'parous':ab,ti OR 'breastfeed*':ab,ti OR 'lactation':ab,ti OR 'lactating':ab,ti OR 'oral contraceptive*':ab,ti OR 'hormone therap*':ab,ti OR 'hormone replacement*':ab,ti OR 'estrogen replacement*':ab,ti OR 'postmenopausal hormon*':ab,ti OR 'age at menopause':ab,ti OR 'menopausal age':ab,ti OR 'menopausal status':ab,ti OR 'height':ab,ti OR 'body mass index':ab,ti OR 'quetelet index':ab,ti OR 'bmi':ab,ti OR 'waist-to-hip':ab,ti OR 'waist hip ratio':ab,ti OR 'physical activit*':ab,ti OR 'alcohol':ab,ti OR 'smoking':ab,ti OR 'family history':ab,ti OR 'benign breast disease':ab,ti OR 'bbd':ab,ti OR 'mammographic densit*':ab,ti OR 'breast densit*':ab,ti |
| #3 | Subtype | 'estrogen receptor'/exp OR 'subtype*':ab,ti OR 'hormone receptor*':ab,ti OR 'estrogen receptor*':ab,ti OR 'oestrogen receptor*':ab,ti OR 'triple negative*':ab,ti OR 'luminal*':ab,ti |
| #4 |  | #1 AND #2 AND #3 |
| #5 | Limits & Filters | #1 AND #2 AND #3 AND ([article]/lim OR [article in press]/lim OR [review]/lim) NOT ([animals]/lim NOT ([animals]/lim AND [humans]/lim)) NOT ('rodent'/exp OR 'dog'/exp OR 'pig'/exp OR 'animal experiment'/exp OR 'animal model'/exp OR 'in vitro study'/exp OR mice:ab,ti OR mouse:ab,ti OR murine:ab,ti OR rat:ab,ti OR rats:ab,ti OR rodent*:ab,ti OR dog:ab,ti OR dogs:ab,ti OR pig:ab,ti OR pigs:ab,ti OR piglet*:ab,ti OR swine*:ab,ti OR porcine*:ab,ti OR 'cell culture*':ab,ti OR 'mcf7':ab,ti OR 't47d':ab,ti OR 'mdamb231':ab,ti) NOT ('letter'/exp OR 'editorial'/exp OR 'news'/exp OR 'conference paper'/exp OR 'retraction notice'/exp OR 'case report'/exp OR 'editorial':ab,ti OR 'conference abstract*':ab,ti OR 'conference paper*':ab,ti OR 'conference review*':ab,ti OR 'conference proceeding*':ab,ti OR 'retraction of publication':ab,ti OR 'retracted publication':ab,ti OR 'corrigenda':ab,ti OR 'corrigendum':ab,ti OR 'errata':ab,ti OR 'erratum':ab,ti OR 'case stud*':ab,ti OR 'retraction notice':ti OR 'protocol':ti OR 'protocols':ti OR 'case report':ti OR 'case series':ti) AND [english]/lim AND [1990-2022]/py |

Database: CINAHL Plus

Platform: EBSCOhost
Date Searched: 7/1/2022
Date: 1/1/1990-7/1/2022

Limit: English | Human/exclude animal studies and non-peer-reviewed

|  | Concept: | Search Strategy: |
| --- | --- | --- |
| #S1 | Breast Cancer | (MH "Breast Neoplasms") OR TI ( "breast neoplasm*" OR "breast cancer*" ) OR AB ( "breast neoplasm*" OR "breast cancer*" ) |
| #S2 | Exposures | ( (MH "Menarche") OR (MH "Menstruation") OR (MH "Breast Feeding") OR (MH "Lactation") OR (MH "Hormone Replacement Therapy") OR (MH "Body Height") OR (MH "Body Mass Index") OR (MH "Waist-Hip Ratio") OR (MH "Alcohol Drinking") OR (MH "Smoking") ) OR TI ( "menarche*" OR "menstrua*" OR "age at first birth" OR "age at first childbirth" OR "parity" OR "parous" OR "breastfeed*" OR "lactation" OR "lactating" OR "oral contraceptive*" OR "hormone therap*" OR "hormone replacement*" OR "estrogen replacement*" OR "progesterone therap*" OR "progesterone replacement*" OR "postmenopausal hormon*" OR "age at menopause" OR "menopausal age" OR "menopausal status" OR "height" OR "Body Mass Index" OR "quetelet index" OR "BMI" OR "waist-to-hip" OR "waist hip ratio" OR "physical activit*" OR "alcohol" OR "smoking" OR "family history" OR "benign breast disease" OR "BBD" OR "mammographic densit*" OR "breast densit*" ) OR AB ( "menarche*" OR "menstrua*" OR "age at first birth" OR "age at first childbirth" OR "parity" OR "parous" OR "breastfeed*" OR "lactation" OR "lactating" OR "oral contraceptive*" OR "hormone therap*" OR "hormone replacement*" OR "estrogen replacement*" OR "progesterone therap*" OR "progesterone replacement*" OR "postmenopausal hormon*" OR "age at menopause" OR "menopausal age" OR "menopausal status" OR "height" OR "Body Mass Index" OR "quetelet index" OR "BMI" OR "waist-to-hip" OR "waist hip ratio" OR "physical activit*" OR "alcohol" OR "smoking" OR "family history" OR "benign breast disease" OR "BBD" OR "mammographic densit*" OR "breast densit*" ) |
| #S3 | Subtype | TI ( "subtype*" OR "hormone receptor*" OR ""estrogen receptor*" OR "oestrogen receptor*" OR "triple negative*" OR "luminal*" ) OR AB ( "subtype*" OR "hormone receptor*" OR ""estrogen receptor*" OR "oestrogen receptor*" OR "triple negative*" OR "luminal*" ) OR (MH "Receptors, Estrogen") |
| #S4 | Limits & Filters | S1 AND S2 AND S3 NOT ((MH "Animals+") OR (MH "Animal Studies")) NOT ("mice" OR "mouse" OR "murine" OR "rat" OR "rats" OR "rodent*" OR "dog" OR "dogs" OR "pig" OR "pigs" OR "piglet*" OR "swine*" OR "porcine*" OR "cell culture*" OR "MCF7" OR "T47D" OR "MDAMB231") NOT ( (MH "Congresses and Conferences") OR (MH "News") OR (MH "Case Studies") OR (MH "Retracted Publication") OR (MH "Retraction of Publication”) )  Limiters - Published Date: 19900101-20220701; Peer Reviewed; English Language; Expanders - Apply equivalent subjects |

Database: Web of Science (Core Collection)

Platform: Clarivate Analytics
Date Searched: 7/1/2022
Date: 1/1/1990-7/1/2022

Limit: English | Human/exclude animal studies and non-peer-reviewed

|  | Concept: | Search Strategy: |
| --- | --- | --- |
| #1 | Breast Cancer | "breast neoplasm*" OR "breast cancer*" (Title) or "breast neoplasm*" OR "breast cancer*" (Abstract) |
| #2 | Exposures | menarche* OR menstrua* OR "age at first birth" OR "age at first childbirth" OR parity OR parous OR breastfeed* OR lactation OR lactating OR "oral contraceptive*" OR "hormone therap*" OR "hormone replacement*" OR "estrogen replacement*" OR "progesterone therap*" OR "progesterone replacement*" OR "postmenopausal hormon*" OR "age at menopause" OR "menopausal age" OR "menopausal status" OR height OR "Body Mass Index" OR "quetelet index" OR "BMI" OR "waist-to-hip" OR "waist hip ratio" OR "physical activit*" OR alcohol OR "smoking" OR "family history" OR "benign breast disease" OR "BBD" OR "mammographic densit*" OR "breast densit*" (Title) or menarche* OR menstrua* OR "age at first birth" OR "age at first childbirth" OR parity OR parous OR breastfeed* OR lactation OR lactating OR "oral contraceptive*" OR "hormone therap*" OR "hormone replacement*" OR "estrogen replacement*" OR "progesterone therap*" OR "progesterone replacement*" OR "postmenopausal hormon*" OR "age at menopause" OR "menopausal age" OR "menopausal status" OR height OR "Body Mass Index" OR "quetelet index" OR "BMI" OR "waist-to-hip" OR "waist hip ratio" OR "physical activit*" OR alcohol OR "smoking" OR "family history" OR "benign breast disease" OR "BBD" OR "mammographic densit*" OR "breast densit*" (Abstract) |
| #3 | Subtype | subtype* OR "hormone receptor*" OR "estrogen receptor*" OR "oestrogen receptor*" OR "triple negative*" OR luminal* (Title) or subtype* OR "hormone receptor*" OR "estrogen receptor*" OR "oestrogen receptor*" OR "triple negative*" OR luminal* (Abstract) |
| #4 |  | DT=(Book OR Book Chapter OR Book Review OR Correction OR Correction, Addition OR Editorial Material OR Item Withdrawal OR Letter OR Meeting Abstract OR Meeting Summary OR News Item OR Note OR Proceedings Paper OR Publication with Expression of Concern OR Retracted Publication OR Retraction OR Withdrawn Publication) |
| #5 |  | animal OR animals OR mice OR mouse OR murine OR "rat" OR "rats" OR rodent* OR rodentia OR dog OR dogs OR pig OR pigs OR piglet* OR swine* OR porcine* OR "cell culture*" OR "MCF7" OR "T47D" OR "MDAMB231" (Topic) or "conference abstract" OR "conference review*" OR "conference proceeding*" OR retracted OR corrigenda OR corrigendum OR errata OR erratum OR "case stud*" OR "retraction notice" OR "protocol" OR "protocols" "case report" OR "case series" (Topic) |
| #6 | Limits & Filters | ((#1 AND #2 AND #3) NOT #4) NOT #5  AND LA=(English)  Timespan: 1990-01-01 to 2022-07-01 (Publication Date) |

Database: Scopus

Platform: Elsevier
Date Searched: 7/1/2022
Date: 1/1/1990-7/1/2022

Limit: English | Human/exclude animal studies and non-peer-reviewed

|  | Concept: | Search Strategy: |
| --- | --- | --- |
| #1 | Breast Cancer AND Exposures AND Subtype Limits | TITLE-ABS ("breast neoplasm*" OR "breast cancer*") AND TITLE-ABS (menarche* OR menstrua* OR "age at first birth" OR "age at first childbirth" OR parity OR parous OR breastfeed* OR lactation OR lactating OR "oral contraceptive*" OR "hormone therap*" OR "hormone replacement*" OR "estrogen replacement*" OR "progesterone therap*" OR "progesterone replacement*" OR "postmenopausal hormon*" OR "age at menopause" OR "menopausal age" OR "menopausal status" OR height OR "Body Mass Index" OR "quetelet index" OR "BMI" OR "waist-to-hip" OR "waist hip ratio" OR "physical activit*" OR alcohol OR "smoking" OR "family history" OR "benign breast disease" OR "BBD" OR "mammographic densit*"OR  "breast densit*") AND TITLE-ABS (subtype* OR "hormone receptor*" OR "estrogen receptor*" OR "oestrogen receptor*" OR "triple negative*" OR luminal*) AND NOT TITLE-ABS (animal OR animals OR mice OR mouse OR murine OR "rat" OR "rats" OR rodent* OR rodentia OR dog OR dogs  OR pig OR pigs OR piglet* OR swine* OR porcine* OR "cell culture*" OR "MCF7" OR "T47D" OR "MDAMB231") AND NOT TITLE-ABS ("conference abstract" OR "conference review*" OR "conference proceeding*" OR retracted OR corrigenda OR corrigendum OR errata OR erratum  OR "case stud*" OR "retraction notice" OR "protocol" OR "protocols" OR "case report" OR "case series") AND LANGUAGE (english) AND PUBYEAR > 1989 AND PUBYEAR < 2023 AND (EXCLUDE (DOCTYPE, "cp") OR EXCLUDE (DOCTYPE "ch") OR EXCLUDE (DOCTYPE, "sh") OR EXCLUDE (DOCTYPE, "le") OR EXCLUDE (DOCTYPE, "ed") OR EXCLUDE (DOCTYPE, "no") OR EXCLUDE (DOCTYPE, "er) OR EXCLUDE (DOCTYPE, "tb") OR EXCLUDE (DOCTYPE, "cr") OR EXCLUDE (DOCTYPE, "bk")) |
